# Deleterious effects of SARS-CoV-2 infection on human pancreatic cells

**DOI:** 10.1101/2021.02.01.21250846

**Authors:** Hanan Shaharuddin, Victoria Wang, Roberta S. Santos, Andrew Gross, Yizhou Wang, Harneet Jawanda, Yi Zhang, Wohaib Hasan, Gustavo Garcia, Vaithilingaraja Arumugaswami, Dhruv Sareen

**Affiliations:** Board of Governors Regenerative Medicine Institute. Cedars-Sinai Medical Center, Los Angeles, CA, USA; Cedars-Sinai Biomanufacturing Center, Cedars-Sinai Medical Center, Los Angeles, CA, USA; Department of Biomedical Sciences, Cedars-Sinai Medical Center, Los Angeles, CA, USA; Genomics Core, Cedars-Sinai Medical Center, Los Angeles, CA, USA; Biobank and Translational research Core, Samuel Oschin Comprehensive Cancer Institute (SOCCI), Cedars-Sinai Medical Center, Los Angeles, CA, USA; Department of Pathology and Laboratory Medicine, Cedars-Sinai Medical Center, Los Angeles, CA, USA; Department of Molecular and Medical Pharmacology, David Geffen School of Medicine, University of California, Los Angeles, Los Angeles, CA 90095, USA; Eli and Edythe Broad Center of Regenerative Medicine and Stem Cell Research, University of California, Los Angeles, Los Angeles, CA 90095, USA; iPSC Core, David and Janet Polak Foundation Stem Cell Core Laboratory. Cedars-Sinai Medical Center, Los Angeles, CA, USA

**Author notes:** Corresponding authors: Dhruv Sareen. Cedars-Sinai Biomanufacturing Center. 8687 Melrose Blvd., Ste. B227, Rm. A252. West Hollywood, CA 90069, USA. Vaithilingaraja Arumugaswami. Department of Molecular and Medical Pharmacology, David Geffen School of Medicine, University of California, Los Angeles, Los Angeles, CA 90095, USA. These authors contributed equally (Hanan Shaharuddin, Victoria Wang, Roberta S. Santos).

**Keywords:** COVID-19, SARS-CoV-2, stem cells, iPSCs, exocrine pancreas, acinar cells, ductal cells, endocrine pancreas, diabetes, pancreatitis, patients

## Abstract

COVID-19 pandemic has infected more than 46 million people worldwide and caused more than 1.2 million deaths. It is transmitted by the Severe Acute Respiratory Syndrome Coronavirus 2 (SARS-CoV-2) and affects the respiratory tract as well as extra-pulmonary systems, including the pancreas, that express the virus entry receptor, Angiotensin-Converting Enzyme 2 (ACE2) receptor. Importantly, the endocrine and exocrine pancreas, the latter composed of ductal and acinar cells, express high levels of ACE2, which correlates to impaired functionality characterized as acute pancreatitis observed in some cases presenting with COVID-19. Since acute pancreatitis is already one of the most frequent gastrointestinal causes of hospitalization in the U.S. and the majority of studies investigating the effects of SARS-CoV-2 on the pancreas are clinical and observational, we utilized human iPSC technology to investigate the potential deleterious effects of SARS-CoV-2 infection on iPSC-derived pancreatic cultures containing endocrine and exocrine cells. Interestingly, SARS-CoV-2 is capable of infecting iPSC-derived pancreatic cells, thus perturbing their normal molecular and cellular phenotypes. The infection increased a key inflammatory cytokine, CXCL12, known to be involved in pancreas dysfunction. Transcriptome analysis of infected pancreatic cultures confirmed that SARS-CoV-2 hijacks the ribosomal machinery in these cells. Notably, the SARS-CoV-2 infectivity of the pancreas is confirmed in post-mortem tissues from COVID-19 patients, which showed co-localization of SARS-CoV-2 in pancreatic endocrine and exocrine cells and increased the expression of some pancreatic ductal stress response genes. Thus, we demonstrate for the first time that SARS-CoV-2 can directly infect human iPSC-derived pancreatic cells with supporting evidence of presence of the virus in post-mortem pancreatic tissue of confirmed COVID-19 human cases. This novel model of iPSC-derived pancreatic cultures will open new avenues for the comprehension of the SARS-CoV-2 infection and potentially establish a platform for endocrine and exocrine pancreas-specific antiviral drug screening.

## INTRODUCTION

The novel coronavirus disease 2019 (COVID-19), caused by SARS-CoV-2 (Severe Acute Respiratory Syndrome Coronavirus-2), initiated in China at the end of 2019 and rapidly escalated to global outbreak, infecting more than 101 million people and resulting in related fatalities in more than 2.1 million by the end of January (JHU Dataset 2021). Although predominantly considered a respiratory disease, nearly one quarter of COVID-19 patients present other symptoms not related to the respiratory tract but to the gastrointestinal (GI) system (Cevik et al., 2020; Liu et al., 2020). SARS-CoV-2 enters host cells through binding of virus Spike protein (S) to ACE2 (Angiotensin-Converting Enzyme 2) receptor and activation of the virus by TMPRSS2 (Transmembrane Protease, serine 2). Interestingly, many other cells besides lung alveolar epithelial cells express ACE2 receptors, including heart, pancreas, GI tract, kidney, testis and other organs (Liu et al., 2020), thus making them a target for the virus. This is supported by studies came demonstrating that virus can affect other tissues, including the heart (Sharma et al., 2020), vascular endothelial cells (Varga et al., 2020), kidney (Pelayo et al., 2020), liver (Shukla and Mohanka, 2020) and the pancreas (Mukherjee et al. 2020; Pinte and Baicus 2020; Tuttolomondo et al. 2020).

The pancreas, specifically the exocrine compartment (acinar and ductal cells), has high expression of ACE2 as analyzed by bulk RNA-seq data (GTEx database) and single-cell RNA-seq of the pancreas (NCBI-GEO database) (Liu et al., 2020). GTEx and TCGA (Genomic Data Commons Data Portal) datasets from human pancreas indicate no differences in ACE2 expression between males or females, as well as between younger (age ≤ 49 years) and older (age ≥ 49 years) populations (Li et al., 2020). In addition, recent findings have shown protein expression of ACE2 and TMPRSS2 in pancreatic cells from healthy humans, especially in the exocrine portion of the pancreas (Coate et al., 2020; Kusmartseva et al., 2020). Furthermore, few clinical reports provide evidence that some patients infected with SARS-CoV-2 who have never presented pancreatic injuries show signs of acute pancreatitis-like symptoms, including elevated levels of serum amylase and lipase, and enlargement of the pancreas as seen in imaging evaluation (Liu et al., 2020). Acute pancreatitis is the major GI cause of hospitalization in the U.S.A., and despite the most common etiologies of this disease being related to gallstones and alcohol abuse, 10% of the cases are caused by infectious microorganisms including viruses (Rawla et al. 2017). Pertinent to the endocrine pancreas, it is now well-established that patients with type I (T1D) and II (T2D) diabetes have a higher risk for COVID-19 associated mortality, especially patients with associated comorbidities such as cardiovascular diseases and renal impairment, as indicated by a study with a large cohort of diabetic patients infected with SARS-CoV-2 (Holman et al., 2020). Diabetic patients (mostly T2D) infected with SARS-CoV-2 presented worse outcomes compared to non-diabetic patients, such as admission to the ICU, mechanical ventilation requirement and higher mortality rates (Apicella et al., 2020). Nevertheless, there is a paucity of knowledge whether and how SARS-CoV-2 can infect and impact the pancreas. This highlights the need for additional investigations to dissect and understand the potential influence of SARS-CoV-2 on human endocrine and exocrine pancreas.

Since primary human pancreatic cells are largely inaccessible and established cell lines do not fully represent human pancreas pathophysiology, we have developed novel methods to generate pancreatic progenitors from human induced pluripotent stem cells (iPSCs), which can be differentiated into endocrine (islet β-cells) and exocrine (acinar and ductal) cells. These cultures contain cells that are representative of human endocrine pancreas expressing endocrine NKX6.1 and C-peptide, exocrine acinar Amylase and Chymotrypsin (CTRC) and exocrine ductal Cytokeratin19 (CK19) and SOX9 cells. We show for the first time that iPSC-derived pancreatic cells including endocrine and exocrine cell types are susceptible to SARS-CoV-2 infection, resulting in morphological perturbations as well as impaired expression of key markers. Importantly, these cellular phenotypes corresponded with inflammatory signatures. Infection of pancreatic tissue was also confirmed in post-mortem pancreatic tissues from COVID-19 patients. Thus, these results suggest the pancreatic cells are susceptible to direct infection by SARS-CoV-2. The iPSC-based model described here provides a valuable novel platform for understanding the pancreas-specific cellular responses to SARS-CoV-2 as well as for antiviral drug development against SARS-CoV-2.

## METHODS

### Ethics Statement and Institutional Approvals

Human cell lines, tissues and histology specimens were obtained or created at Cedars-Sinai under the auspices of the Cedars-Sinai Medical Center Institutional Review Board (IRB) approved protocols. Specifically, the iPSC cell lines and differentiation protocols in the present study were carried out in accordance with the guidelines approved by Stem Cell Research Oversight committee (SCRO) and IRB, under the auspices of IRB-SCRO Protocols Pro00032834 (iPSC Core Repository and Stem Cell Program) and Pro00036896 (Sareen Stem Cell Program). Infections of iPSC-derived cells were performed under the auspices of UCLA’s Stem Cell Oversight Committee under protocol #2020-004-01 and UCLA Biosafety Committee protocol BUA-2020-015-004-A. Post-mortem tissues were collected by Cedars-Sinai’s Biobank and Translational Research core in accordance with protocol Pro00036514.

### Human iPSC culture

The induced pluripotent stem cell (iPSC) lines utilized in this study were generated from healthy volunteers at the iPSC Core at Cedars-Sinai Medical Center from the peripheral blood mononuclear cells (PMBCs) utilizing non-integrating oriP/EBNA1-based episomal plasmid vectors, as described in(Rajamani et al., 2018). This approach results in highly cytogenetically stable iPSCs as tested by G-band karyotyping. All undifferentiated iPSCs were maintained in mTeSR^+^ media (StemCell Technologies, Cat 05825) onto BD Matrigel™ matrix-coated plates.

### Generation of iPSC-differentiated Pancreatic Progenitors

iPSCs were single-cell dissociated using Accutase and plated onto Matrigel-coated plates at a density of 300,000 cells/cm^2^ using mTeSR^+^ and 10 µM Rho kinase Inhibitor (Stem Cell Tech). The following day, cells were directed into Definitive Endoderm (DE) using Phase I medium, which was composed of base medium MCDB 131 (Fisher Sci) supplemented with 100 ng/ml Activin A (R&D), 2 µM CHIR99021 (Stemgent), and 10 µM Rho kinase Inhibitor (Stem Cell Tech.) for 1 day. For the next two days, the same base medium was used, but supplemented instead with 100 ng/mL Activin A and 5 ng/mL FGF2 (Peprotech). Following this phase, cells were directed to form Posterior Foregut (PFG) using Phase II medium, which was composed of the same base medium as Phase I but supplemented with 50 ng/mL FGF10 (Peprotech), 0.25 µM CHIR99021 and 50 ng/ml Noggin (Peprotech), for 2 days. To reach a Pancreatic Progenitor (PP) stage, cells were fed with Phase III medium, which was composed of DMEM supplemented with 50 ng/mL Noggin, 50 ng/mL FGF10, 2 µM Retinoic Acid (Sigma), and 0.25 µM SANT1 (Sigma), for four days. More details about base medium formulation are described at **Suppl. Table 1**. This protocol was based on a published protocol to differentiate iPSCs into Pancreatic Progenitors (Memon et al., 2018).

### Generation of iPSC-differentiated Pancreatic Endocrine (iPan^ENDO^), Acinar (iPan^EXO^ Acinar) and Ductal (iPan^EXO^ Ductal) cells

To generate iPan^EXO^ Ductal cells, on Day 7 of differentiation, cells were single cell dissociated and seeded at 103.1k cells/cm^2^ in Phase III medium supplemented with 10 µM Rho kinase Inhibitor on a Matrigel-coated plate. From Day 8 to Day 26, cells were grown in iPan^EXO^ Ductal phase media, which consisted of Phase III base medium supplemented with 25 ng/ml FGF10, 50 ng/ml EGF (Peprotech), and 25 ng/ml sDLL-1 (Peprotech). To generate iPan^ENDO^ and iPan^EXO^ Acinar cells, based on Hohwieler et al. (2017) protocol, after 4 days of Phase III media, PPs were fed with Phase III base medium supplemented with with 20 ng/mL FGF10, 1µM XXI (Sigma Aldrich), 50 ng/mL Noggin, 10 mM Nicotinamide (Sigma-Aldrich), and 25 ng/mL Wnt3a (Peprotech) for seven days, daily feeding (Hohwieler et al., 2017). More details about base medium formulation are described at **Suppl. Table 1**.

### Generation and Maintenance of iPan^EXO^ Organoid Cultures

On the last day of Phase III, PPs were roughly dissociated via scraping and trituration. They were centrifuged at 170 x G for 3 minutes, and then resuspended with a solution composed of Phase III medium and Matrigel at a 1:4 ratio (1 of medium and 4 of Matrigel). 30 µL of this solution with cells was plated into each well of a 96 round bottom plate and then incubated at 37 °C for 20 minutes, before the plate was flipped upside down for 10 minutes. 100 µL of Phase III base medium supplemented with 20 ng/mL FGF10, 50 ng/mL Noggin, 10 mM Nicotinamide, and 25 ng/mL Wnt3a was then added to the cells. Cells were fed every other day with the same media until Day 57 or when organoids contained lumen and were at least 150 µm large.

### SARS-CoV-2 stock

SARS-CoV-2, isolate USA-WA1/2020, was obtained from the Biodefense and Emerging Infections (BEI) Resources of the National Institute of Allergy and Infectious Diseases (NIAID). Importantly, all studies involving SARS-CoV-2 infection of iPSC-derived pancreatic cells were conducted within a Biosafety Level 3 high containment facility at UCLA. SARS-CoV-2 was passed once in Vero-E6 cells and viral stocks were aliquoted and stored at – 80 °C. Virus titer was measured in Vero-E6 cells by TCID50 assay. Vero-E6 cells were cultured in DMEM growth media containing 10% FBS, 2 mM glutamine, pen/step, and 10 mM HEPES. Cells were incubated at 37 °C with 5% CO_2_.

### SARS-CoV-2 infection of iPSC-derived pancreatic (iPan) cultures

SARS-CoV-2 viral inoculum (MOI of 0.05 and 0.1) was prepared using acinar or ductal cell specific media. Human iPSCs were differentiated into iPSC-derived pancreatic (iPan) cultures containing iPan^ENDO^, iPan^EXO^ Acinar and iPan^EXO^ Ductal cells in 96-well or 24-well plates before infection as detailed above, and the culture media at Day 26 of differentiation for iPan^EXO^ Ductal and Day 16 for iPan^EXO^ Acinar were replaced with 100 μl of prepared inoculum. For mock infection, cell type specific media (100 μl/well) alone was added. The inoculated plates were incubated for 1 hour at 37 °C with 5% CO_2_. At the end of incubation, the inoculum was replaced with fresh iPan^EXO^ Acinar or iPan^EXO^ Ductal culture medium. Cells remained at 37 °C with 5% CO_2_ for 24 hours (Day 1) or 72 hours (Day 3) before analysis. All studies involving active SARS-CoV-2 infection of iPSC-derived pancreatic cell cultures were conducted within a Biosafety Level 3 facility at University of California in Los Angeles (UCLA), CA, USA.

### Immunofluorescence and imaging of cells

Cells subjected to immunofluorescence were fixed with 4% paraformaldehyde (PFA) in phosphate-buffered saline (PBS) for 20 minutes and subsequently washed with PBS. Fixed cells were then permeabilized and blocked for 1 hour in a blocking buffer containing PBS with 10% donkey serum (Millipore) and 0.1% Triton-X (Bio-Rad). Primary antibodies were diluted in the blocking buffer and added to the cells overnight at 4 °C. The following primary antibodies and dilutions were used: SARS Coronavirus (1:400, NR-10361, BEI Resources) SARS-CoV-2 Spike S1 (1:100, 40150-R007, SinoBiological), eCAD (1:100, AF68, R&D Systems), MIST1 (1:100, MA1517, Invitrogen), CK19 (1:100, PIMA512663, Thermofisher), SOX9 (1:250, AB5535,Millipore), ACE2 (1:100, AB15348, abcam), Chymotrypsin (1:100, MAB1476, Millipore), and Amylase (1:100, A8273, Sigma). After washing using PBS with 0.1% Tween-20 (ThermoFisher), cells were incubated with appropriate species-specific Alexa Fluor-conjugated secondary antibodies (ThermoFisher) diluted in a blocking buffer (1:1,000) for 1 hour at room temperature. After washing in PBS with 0.1% Tween-20, cells were incubated in Hoechst 33342 diluted in PBS (1:2,500) for 15 min. Immunofluorescence images were visualized using appropriate fluorescent filters using ImageXpress Micro XLS (Molecular Devices) and analyzed using ImageJ Software. Image quantification was performed using CellProfiler Software (v3.1.9).

### Real-Time qPCR

Relative gene expression was quantified using RT-qPCR. For this, cells were washed with PBS to remove any possible remaining viral inoculum. and the total RNA was isolated with RLT (Qiagen). RNA was then extracted with RNeasy Micro Kit (Qiagen) according to the manufacturer’s instructions. The concentration of RNA was determined by spectrophotometric analysis (Qubit 4 Fluorometer, ThermoFisher) and the purity with NanoDrop (ThermoFisher); all samples had a A_260/280_ ratio around 2.0 (Desjardins and Conklin, 2010). After, RNA (1 μg) was reverse transcribed to cDNA with oligo(dT) using the High Capacity cDNA Reverse Transcription kit (ThermoFisher). Real-time qPCR was performed in triplicates using SsoAdvanced Universal SYBR Green Supermix (Biorad) and specific primer sequences to each gene (**Suppl. Table 2**),on a CFX384 Real Time system (Bio-Rad). Human *RPL13* was used as the reference gene and relative expression was determined using 2^-ΔΔ^ Ct method.

### Post-mortem Human pancreatic tissues

#### a) Real-Time qPCR from snap-frozen tissues

Post-mortem pancreatic samples were isolated from the head of the pancreas (preferably) from patients that were infected with SARS-CoV-2 and passed from complications related to the disease (*COVID-19 patients*), or patients that were not infected with SARS-CoV-2 and passed from complications not related to it (*Control patients*). These samples were isolated 1-3 days after the patient’s death and were snap-frozen in liquid nitrogen. Samples were stored for longer in -80 °C before used for RNA extraction. For RNA isolation, Trizol (Thermo) was used and for RNA extraction, RNeasy Mini Kit (Qiagen) was used according to the manufacturer’s instructions. The concentration of RNA was determined by spectrophotometric analysis (Qubit 4 Fluorometer) and the purity with NanoDrop, as described above; all samples had a A_260/280_ ratio around 2.0. After, RNA (2 μg) was reverse transcribed to cDNA with oligo(dT) using the High Capacity cDNA Reverse Transcription kit. Real-time qPCR was performed in triplicates using SsoAdvanced Universal SYBR Green Supermix (Bio-Rad) and specific primer sequences to each gene can be found at Suppl. Table 3. qPCR was performed on a CFX384 Real Time system. Human *β-ACTIN* was used as the reference gene and relative expression was determined using 2^-ΔΔ^ Ct method.

#### b) Immunohistochemistry from paraffin embedded tissues

Pancreatic tissues were fixed in 10%formalin and then paraffin embedded. Blocks were sectioned at 4 µm thickness and mounted at microscope slides (Superfrost Plus microscope slides, Fisher). Slides were washed 2x in Xylene for 10 minutes each. This was followed by 2x 5-minute washes with 100% ethyl alcohol, 1x 3-minute wash with 95% ethyl alcohol, and 1x 3 minute wash with 75% alcohol for rehydration. They were then washed 3x with PBS for 5 minutes before beginning antigen retrieval. Samples were submerged in 10mM pH 6.0 sodium citrate Buffer, and then microwaved for 10 minutes at 80% power. Once cooled at room temperature for one hour, they were washed 3x for 5 minutes each with PBS. Samples were blocked for 2 hours in the same blocking buffer as mentioned above (PBS with 10% donkey serum and 0.1% Triton-X). Samples were incubated 4 °C overnight with primary antibodies in the same concentrations as mentioned above. The following day, they were washed 3x for 10 minutes each in PBS, incubated one hour at room temperature with the secondary antibodies at concentrations of 1:1000, and then finally washed 3x for 10 minutes each before mounted with ProLong™ Gold Antifade Mountant with DAPI (Invitrogen).

### RNA Sequencing (RNASeq)

#### a) RNA extraction and sequencing

RNA was extracted from pelleted cells using RNeasy Micro Kit (Qiagen) and was prepared for sequencing with the Illumina TruSeq Stranded mRNA library preparation kit (Illumina, San Diego, CA) by the Cedars-Sinai Applied Genomics, Computation, and Translational Core. Concentration and quality of RNA was assessed on a Qubit fluorometer (ThermoFisher Scientific, Waltham, MA) and 2100 Bioanalyzer (Agilent Technologies, Santa Clara, CA) respectively. Complementary DNA was reverse transcribed using Invitrogen’s Reverse Transcriptase kit (Carlsbad, CA) and converted into double-stranded DNA (dsDNA). The dsDNA was then enriched using PCR before purification with Agencourt AMPure XP beads (Beckman Coulter, Brea, CA). The enriched purified DNA was then quantified and resolved via Qubit and Bioanalyzer. Sample libraries were then multiplexed and sequenced on Illumina’s NextSeq 500 platform (San Diego, CA) using 75 bp single-end sequencing.

#### b) Analysis

Raw reads were quantified by mapping them with the STAR aligner (version 2.5.0) (Dobin et al., 2013) / RSEM (version 1.2.25) (Li and Dewey, 2011) to the GRCh38 human reference transcriptome based on human GENCODE version 33 (www.gencodegenes.org) as well as the GenBank: MT246667.1 SARS-CoV-2 reference viral genome. Expression tables were post-processed with a custom R script that filtered out non-coding RNA based on the transcript biotype assigned by biomaRt. Initial analyses were performed on the BioJupies platform. Principal Component Analysis was then performed in R using the prcomp package. Differential expression tables were calculated in R using the DESeq2 package. Differential expression tables were used to plot heatmaps and volcano plots in ggplot. Enrichment analysis was performed by uploading the top 500 upregulated and downregulated transcripts to the Enrichr gene enrichment analysis portal (Chen et al., 2013; Kuleshov et al., 2016). Enrichment results of interest (including Gene Ontology enrichment and COVID-related gene enrichment) were exported from Enrichr as text files and imported into R for plotting using ggplot. All analysis and plotting scripts in R are available on github.com at github.com/Sareen-Lab/COVID. Gene transcript tables as well as the originalFASTQ files are available through NCBI’s GEO database at accession number GSE95243.

### Statistical analyses

Data are presented as mean ± standard error of the mean (SEM). Statistical significance between groups was determined by One-way ANOVA followed by Dunnet post-test. Two-tailed paired Student’s test was used as appropriate. *P* values <.05 were considered statistically significant. Statistical analyses and graphs were generated using GraphPad Prism 7 for Windows Software (GraphPad Software).

## RESULTS

### iPSC-derived pancreatic cells exhibit ACE2 and TMPRSS2 expression

Human iPSCs were differentiated into pancreatic adherent (2D) and organoid (3D) cultures. Protocols for differentiation after the pancreatic progenitor stage were directed to bias the cell fate of the cultures containing either exocrine acinar cells (iPan^EXO^ Acinar) or exocrine ductal cells (iPan^EXO^ Ductal). iPan^EXO^ Acinar cultures express some markers that are characteristic of mature acinar cultures such as cytoplasmic staining of digestive enzymes Amylase (AMY) and Chymotrypsin C (CTRC), and greater staining of nuclear transcription factor MIST1, which is characteristic of acinar cells that are not fully mature. In our cultures, cells that are AMY^-^ are MIST1^+^, which indicates we have mixed cultures of acinar cells containing cells more mature and some less mature. Acinar cultures also contain some C-peptide expressing endocrine islet β-cells (iPan^ENDO^). iPan^EXO^ Ductal cultures express cytoskeletal staining of Cytokeratin 19 (CK19). Interestingly, both AMY^+^ iPan^EXO^ Acinar and CK19^+^ iPan^EXO^ Ductal cultures exhibit ACE2 protein expression (**Figure 1A-C**), which is now a confirmed entry point receptor for SARS-CoV-2. Notably, while coexpression is seen, not all AMY^+^ and C-peptide^+^ cells express ACE2, and this seems partially to be consistent with current literature, which shows lower ACE2 expression in endocrine cells (Coate et al., 2020; Kusmartseva et al., 2020). *ACE2* and *TMPRSS2* gene expression was also measured in pluripotent iPSC stage cells, iPSC-derived pancreatic cultures, human cadaveric pancreatic tissue, and human pancreatic duct epithelium cell line, H6C7. Relative to iPSCs, iPan^EXO^ Acinar and iPan^EXO^ Ductal cultures contain significantly higher levels of *ACE2*. There is also a significantly higher expression of *TMPRSS2* in iPan^EXO^ Acinar and human acinar tissues, but no detection of this gene in H6C7 cell line and low detection in iPan^EXO^ Ductal cells (**Figure 1D**). Differentiated iPan^EXO^ 3D organoids also show co-localization of ACE2 protein along with acinar and ductal markers (**Suppl. Figure 1**).

**Figure 1.**
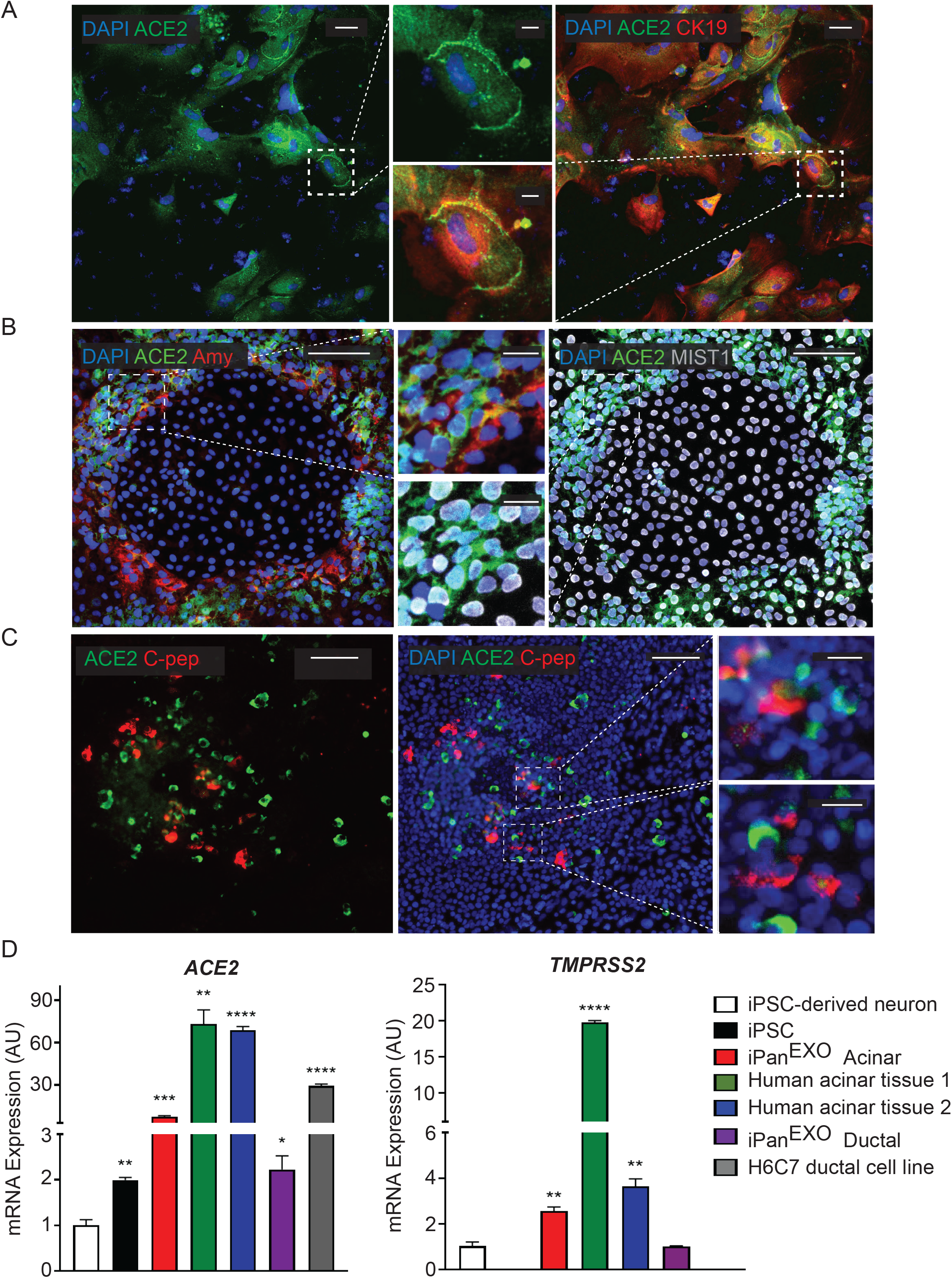
Expression of ACE2 and TMPRSS2 in exocrine pancreatic cells. **A**. iPan^EXO^ Ductal cells CK19 (red) exhibiting ACE2 (green) expression. **B**. iPan^EXO^ Acinar cells Amylase (red), MIST1 (gray) exhibit ACE2 (green) expression. **C**. iPan^EXO^ Acinar and iPan^ENDO^ cultures contain some endocrine C-peptide expressing cells, which also co-stain with ACE2. **D**. iPSC-derived pancreatic exocrine cells as well as human acinar tissues and human ductal cell line H6C7 express *ACE2* and *TMPRSS2*. Data is shown as mean ± SEM with statistical significance determined by unpaired two-tailed t-test. *p<0.05, **p <0.01, ***p<0.001, ****p<0.0001. Scale bar represents 100 µm, and 20 µm for zoomed panels adjacent to main images.

### SARS-CoV-2 can directly infect iPan^EXO^ Ductal cultures and elicit abnormal cellular phenotypes

After 26 days of pancreatic ductal differentiation starting from iPSCs, cultures were infected with SARS-CoV-2 at a multiplicity of infection (MOI) 0.05 and 0.1, and cells were collected for analysis after one day (Day 1) or after three days (Day 3) (**Figure 2A**). These MOIs allowed for establishing de novo infection and active viral replication for the viral kinetics studies. The mock condition was treated with ductal media with no virus. MOI 0.05 infection shows 4% and 19% of cells infected with SARS-CoV-2 on Day 1 and Day 3 respectively, while MOI 0.1 infection shows 10% and 19% of infected cells, respectively **(Figure 2B)**, showing viral infectivity increased from Day 1 to Day 3 in both MOIs. Although more cells got infected from Day 1 to Day 3, less cells were observed in culture comparing infected cultures with uninfected cultures, which suggests that the virus possibly decreased cell viability (data not shown). As expected, the mock condition has negative SARS-CoV-2-Spike S1 staining (**Figure 2B**). RT-qPCR for SARS-CoV-2 nucleocapsid gene *N1* shows SARS-CoV-2 mRNA production increased in the infected conditions compared to mock condition as seen in **Figure 2C**.

**Figure 2.**
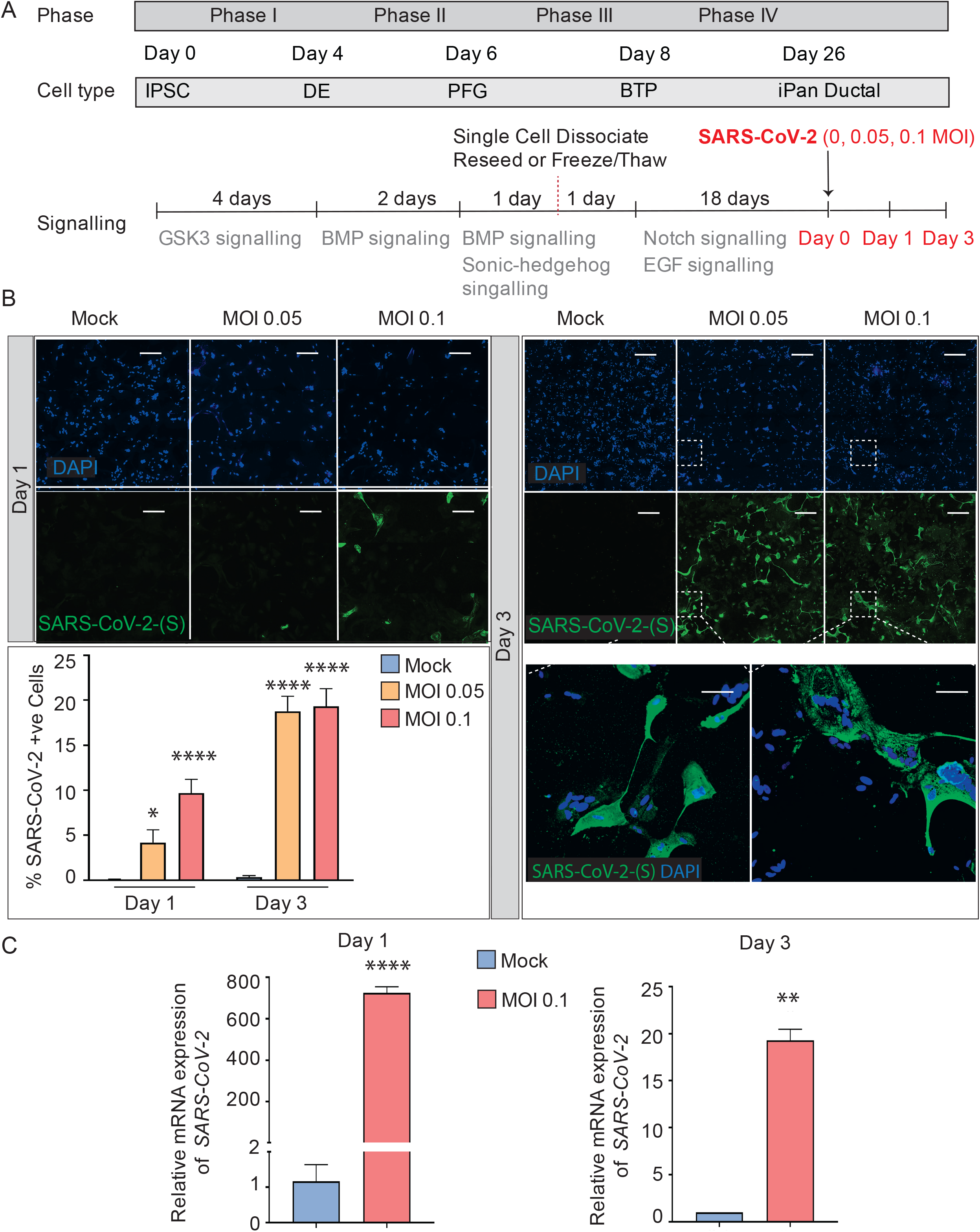
SARS-CoV-2 can infect iPan^EXO^ Ductal Cells. **A**. Differentiation scheme of pancreatic ductal cells and infection of SARS-CoV-2 on Day 26. **B**. Immunocytochemistry staining of SARS-(S) at Day 1 and Day 3 (Scale bar is 200 µm). **C**. RT-qPCR showing upregulation of *SARS-CoV-2 N1* from mock to infected cells at different days. Data is shown as mean ± SEM with statistical significance determined by unpaired two-tailed t-test. *p<0.05, **p <0.01, ****p<0.0001. Scale bar represents 200 µm.

SOX9 is a well-established nuclear transcription factor involved in determining ductal specification from pancreatic progenitors. Interestingly, SARS-CoV-2 positive cells in the infected iPan^EXO^ Ductal cultures at both MOIs show a more enlarged cellular morphology as observed by CK19 and SOX9 staining pattern **(Figure 3A)** and abnormal cytoplasmic SOX9 localization instead of the typical nuclear localization observed in normal ductal cells as evident in the mock condition and uninfected cells **(Figure 3B)**. In fact, we observe a higher number of ductal cells with mislocalization of SOX9 in the cytoplasm of infected cells at both Day 1 and Day 3 compared with mock cells, and both MOIs seem to have a similar pattern of mislocalization (no statistical difference between MOIs in both Day 1 and Day 3) **(Figure 3C)**. Meanwhile, CK19 is present across all treatments and timepoints, with no obvious differences in staining localization of the infected population. ACE2 expression is observed in Day 3 cells with no distinct differences between mock and MOI 0.05 and 0.1 conditions **(Suppl. Figure 2)**.

**Figure 3.**
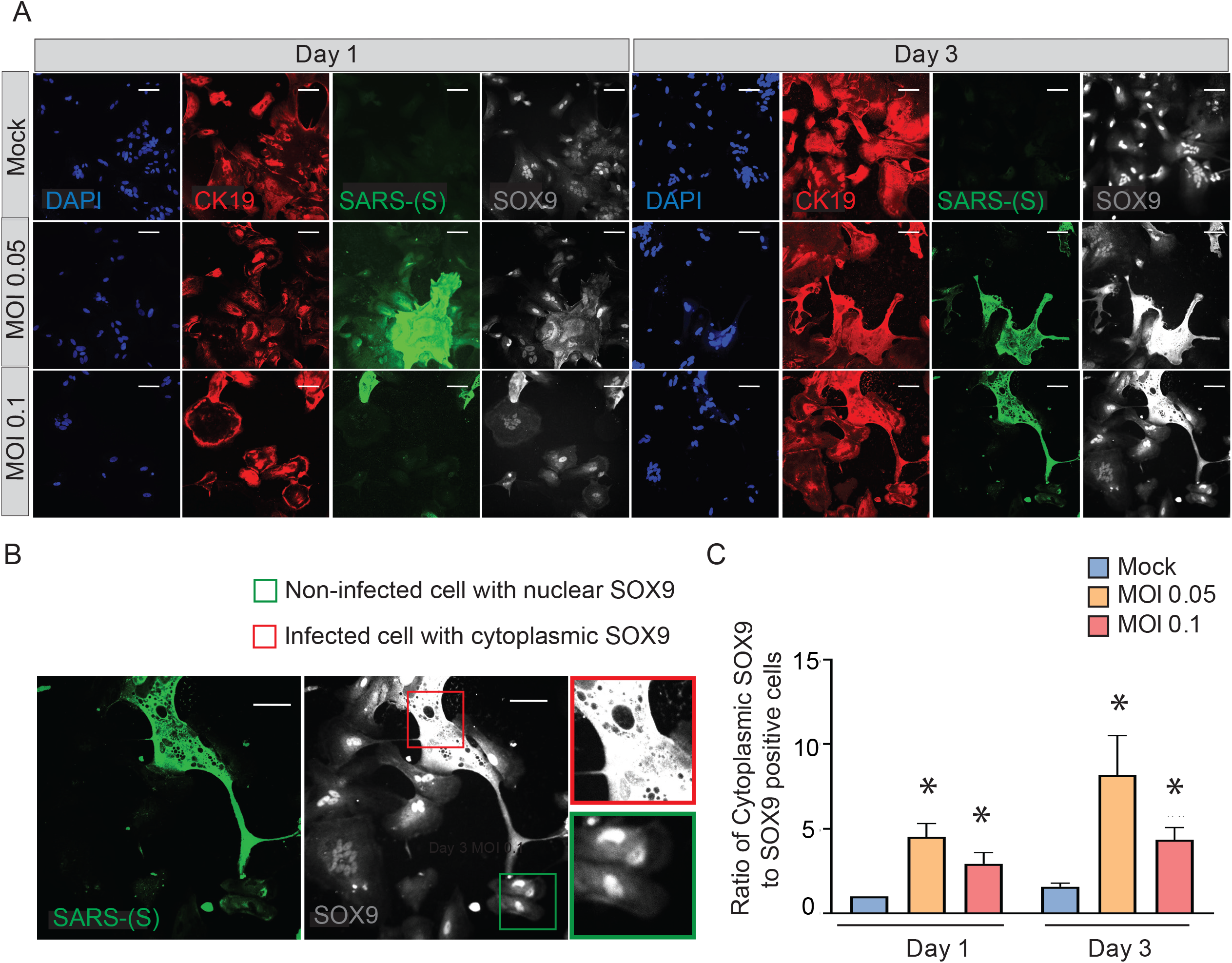
SARS-CoV-2 elicits abnormal cellular phenotypes in iPan^EXO^ Ductal cultures. **A**. Day 1 and Day 3 of post infected iPan^EXO^ Ductal cells. Immunocytochemistry staining of ductal cell markers CK19 (red), SOX9 (gray), and SARS-CoV-2 infected cells (green) on Day 1 and Day 3 with varying titers of virus (Mock, MOI 0.05, and 0.1). **B**. SOX9 translocation in infected cells. (Scale bar is 200 µm). **C**. Histogram showing the ratio of cells with mis localized cytoplasmic SOX9 over total nuclear SOX9 positive cells in the culture, comparing mock vs SARS-CoV-2 infected ductal cultures at MOI 0.05 and 0.1. Data is shown as mean ± SEM with statistical significance determined by unpaired two-tailed t-test. *p<0.05, **p <0.01. Scale bar represents 200 µm. Zoomed panels adjacent to main images are 2.3x larger.

### SARS-CoV-2 can infect iPan^EXO^ Acinar and iPan^ENDO^ cells resulting in activation of proinflammatory genes

iPan^EXO^ Acinar cultures containing predominantly Chymotrypsin C (CTRC) and Amylase (AMY) positive cells and few iPan^ENDO^ C-peptide positive islet β-cells were differentiated for 16 days before being infected with SARS-CoV-2 at a MOI of 0.1. A separate mock condition was cultured in parallel and not infected. Like the iPan^EXO^ Ductal cultures, infected and uninfected cultures were fixed or lysed after one day or three days of infection **(Figure 4A)**. CTRC-positive cells, which have a granular morphology that likely reflects zymogen granule formation, showed direct infection by SARS-CoV-2 **(Figure 4B)**. Interestingly, a subpopulation of SARS-CoV-2 positive cells in these cultures were also C-peptide positive **(Suppl. Figure 3)**. On Day 1 post-infection, 0.63% of total cells were infected, which increased to 1.12% by Day 3. For both days, the difference in percentage of infected cells was statistically significant compared to mock condition **(Figure 4C)**, which was similarly seen on mRNA level by RT-qPCR for SARS-CoV-2 nucleocapsid gene *N1*, showing high mRNA production of SARS-CoV-2 in the infected conditions **(Figure 4D)**.

**Figure 4.**
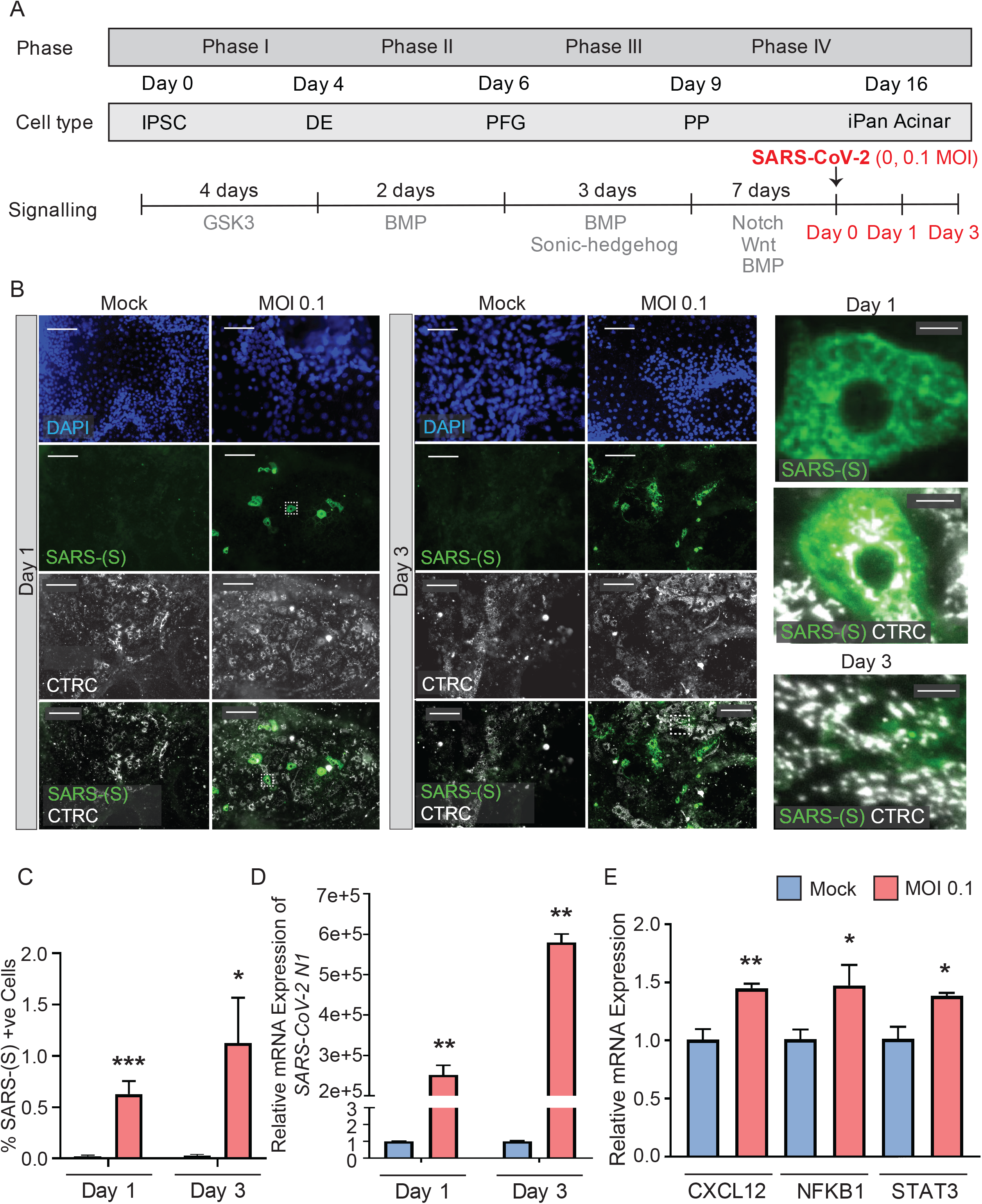
iPan^EXO^ Acinar cultures can be infected with SARS-CoV-2 and result in upregulation of some proinflammatory genes. **A**. Differentiation and infection timeline for the iPan^EXO^ Acinar cells. Cells were grown 16 days before being infected. Cells were fixed or lysed on the first and third days of infection. **B**. Immunocytochemistry staining of SARS-(S) on Day 1 and 3 of mock and infected cells. **C**. Quantification of immunocytochemistry images shows significant increase in SARS-(S) positive cells in the population treated with the virus at MOI 0.1. **D**. qPCR shows upregulation of *SARS-CoV-2 N1* in infected cells. **E and F**. qPCR of inflammation markers *CXCL12, NFKB1*, and *STAT3* show significant upregulation between mock and infected cells on day 3 of infection. Data is shown as mean ± SEM with statistical significance determined by unpaired two-tailed t-test. *p<0.05, **p<0.01, ***p<0.001. Scale bar represents 130 µm, and 10 µm for zoomed panels adjacent to main images.

To investigate whether SARS-CoV-2 infection perturbed the inflammatory pathway, multiple genes known to be associated with pancreatitis-related inflammation were assessed using RT-qPCR. Among those, the genes *CXCL12, NFKB1*, and *STAT3* showed significant upregulation at Day 3 in infected cells compared to mock condition (**Figure 4E**), while no significant change was observed on infected cells by Day 1 compared to the mock condition **(Suppl. Figure 4)**. No significant change in expression was seen on other inflammatory markers, such as *IL1B* and *TNFA*, on both days **(Suppl. Figure 4)**.

### Transcriptional analysis of SARS-CoV-2 infected iPSC-derived pancreatic cultures demonstrates active viral replication and pancreas-specific COVID-19 associated disease signatures

The iPan cultures containing iPan^EXO^ Acinar and iPan^ENDO^ cells infected with 0.1 MOI SARS-CoV-2 were harvested on Day 1 and Day 3 post-infection for transcriptomic analysis after mRNA sequencing, as well as cells that were not infected (mock conditions). After mapping genomic reads in infected cultures, mapped reads were detected from the SARS-CoV-2 genome that confirm active SARS-CoV-2 viral replication within infected cultures (**Suppl. Table 3**). Principal component analysis (PCA) of Day 1 and Day 3 infected cultures demonstrated that 37.9% and 41.1% of the variance in the gene expression differences could be attributed to principal component 1 (PC1) in Day 1 and Day 3 infected cultures, respectively (**Figure 5A**). Both PCA and the heatmaps of differentially expressed genes clearly demonstrated transcriptomic clustering of samples in either the mock or infected condition in both days (**Figure 5B**). Upon further interrogation of the Day 3 data, SARS-CoV-2 infection induced significant pancreas-specific gene expression changes within iPSC-derived pancreatic cultures including upregulation of *PDX1, INS, GCG, CHGA, CFTR, IGFBP7* and *GHRL* and downregulation of genes involved in cellular secretory pathway such as *GOLGA8A* and *GOLGA8B*. Significant changes in expression of chemokine and immunomodulatory genes were detected and most were upregulated such as *PTGES, MIF, CCR7, CXCL6* and *CXCL12*, which encodes immune cytokines known to be transcriptionally upregulated during SARS-CoV infection. Similarly, genes reflecting pathogenic interaction and antiviral responses in host cells such as *THOC1, TRIM28, CD37, TRAF3IP1, DDX17, NCBP3, HYAL2*, were also perturbed (**Suppl Table 4**). Gene enrichment pathway analysis confirmed significant upregulation of biological processes transcriptional pathways related to signal recognition particle (SRP)-dependent translation and targeting of proteins to the endoplasmic reticulum (ER) membrane, viral transcription and viral gene expression (**Figure 5D**), consisting of mainly ribosomal complex large and small subunit genes. This is consistent with the idea that SARS-CoV-2 like many other viruses requires recruitment of a variety of host cell factors including ribosomal proteins to participate in viral protein biosynthesis, in order to survive, accumulate and propagate in the pancreatic cells. The most significant cellular components that are downregulated include nuclear body, nuclear specks and nucleoplasm (**Figure 5E**), which is also consistent with the downregulation of the specific nuclear pore complex (NPC) genes such as *NPIPB3, 4, 5* (**Figure 5C**). Numerous viral pathogens have evolved different mechanisms to hijack the NPC in order to regulate trafficking of viral proteins, genomes and even capsids into and out of the nucleus thus promoting virus replication (Le Sage and Mouland, 2013). As expected, the cytosolic ribosomal units are the most significantly upregulated cellular component (**Figure 5E**). Notably, the virus perturbation signatures with SARS-CoV-2 in iPSC-derived pancreatic cells were similar to responses by previously published SARS-CoV, SARS-dORF6 and SARS-BatSRBD infection of human lung alveolar epithelium cells (**Figure 5F**) (Mitchell et al., 2013). Upon comparing COVID-19 associated transcripts published and available in the Enrichr database, similar signatures were significantly perturbed in SARS-CoV-2-infected iPan^EXO^ Acinar, consistent with prior reports examining viral infection in pancreas (**Figure 5G**). Taken together, these results indicate that SARS-CoV-2 infection induces significant transcriptional changes within iPSC-derived pancreatic cultures.

**Figure 5.**
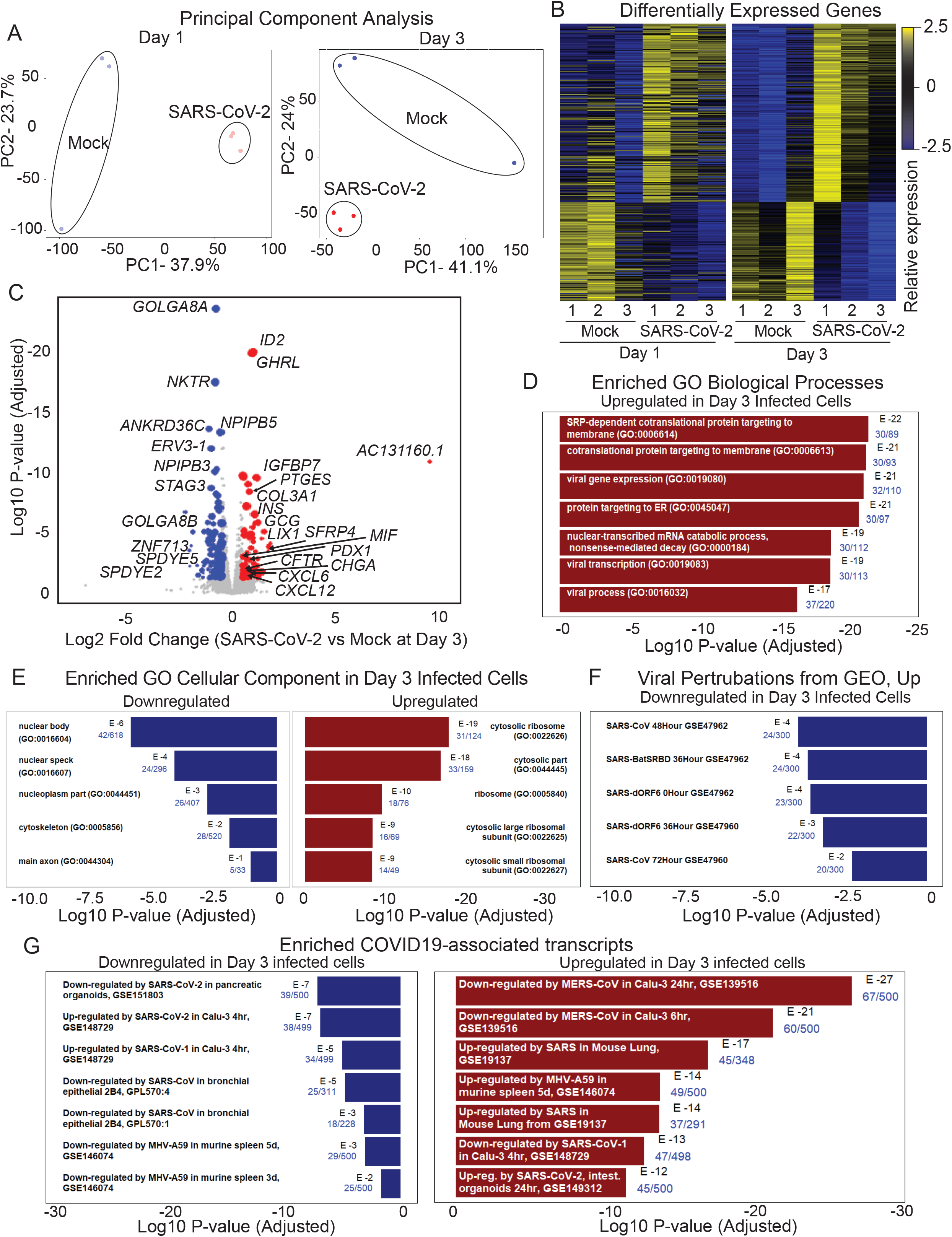
RNAseq analysis of infected iPan^EXO^ cells on days 1 and 3 post infection. **A**. Principal Component Analysis of infected and mock samples collected on day 1 and day 3, respectively. **B**. Differentially Expressed Genes Heat map showing the relative expression levels of transcripts differentially expressed with adjusted p-values less than 0.01. **C**. Volcano plot of the log10 adjusted p-value of each expressed transcript on day 3 versus the log2 fold change. Transcripts that did not demonstrate differential expression with an adjusted p-value of less than 0.05 and a log2 fold-change in either direction greater than 0.5 are plotted in grey. Those that did were sized proportionally to their mean expression level, and genes of interest have been labeled. **D**. Gene Ontology terms in the Biological Process database were plotted based on the probability of the observed enrichment. The 500 most upregulated and the top 500 most downregulated transcripts were submitted to Enrichr. The fraction of genes in the gene set which were present in the 500 transcripts under analysis are listed in blue. **E**. Enrichment scores for the 500 most downregulated transcripts on day 3 when compared to the Gene Expression Omnibus’ gene sets of genes upregulated in response to viral perturbation(Barrett et al., 2013) **F**. Enrichment scores for the most upregulated and downregulated transcripts on day 3 within the COVID-19-related Drug and Gene Set Library (Kuleshov et al., 2020).

### Post-mortem human pancreatic tissues from *COVID-19 patients* show infectivity and perturbed expression of pancreatic genes

Post-mortem human pancreatic samples were obtained from individuals who succumbed from complications related to COVID-19 infection (COVID-19 patients) or from those who were not infected by SARS-CoV-2 and were deceased due to complications unrelated to COVID-19 (Control patients). Samples were processed for immunohistochemistry of SARS-CoV-2-Spike S1 and pancreatic markers, or processed for analysis of changes in gene expression. As shown in **Figure 6 A-C**, SARS-CoV-2-S was detected in pancreas of COVID-19 patients, demonstrating the susceptibility of the human pancreas to the virus. To better understand which cell types were infected by SARS-CoV-2 in the pancreas, cells were stained for specific endocrine and exocrine markers, such as Amylase and Chymotrypsin (CTRC) for acinar cells, Cytokeratin 19 (CK19) for ductal cells, and C-peptide for endocrine islet β-cells. Interestingly, many of the pancreatic cell types tested here co-localized with SARS-CoV-2-S. The virus was present in the majority of the CTRC^+^ and some clusters of Amylase^+^ cells **(Figure 6A-B)**. However, as for the ductal population, SARS-CoV-2-S was not detected inCK19^+^ cells **(Suppl. Figure 5)**. From the endocrine side, interestingly, SARS-CoV-2 staining was frequently concentrated in C-Peptide^+^ islet clusters **(Figure 6E)**.

**Figure 6.**
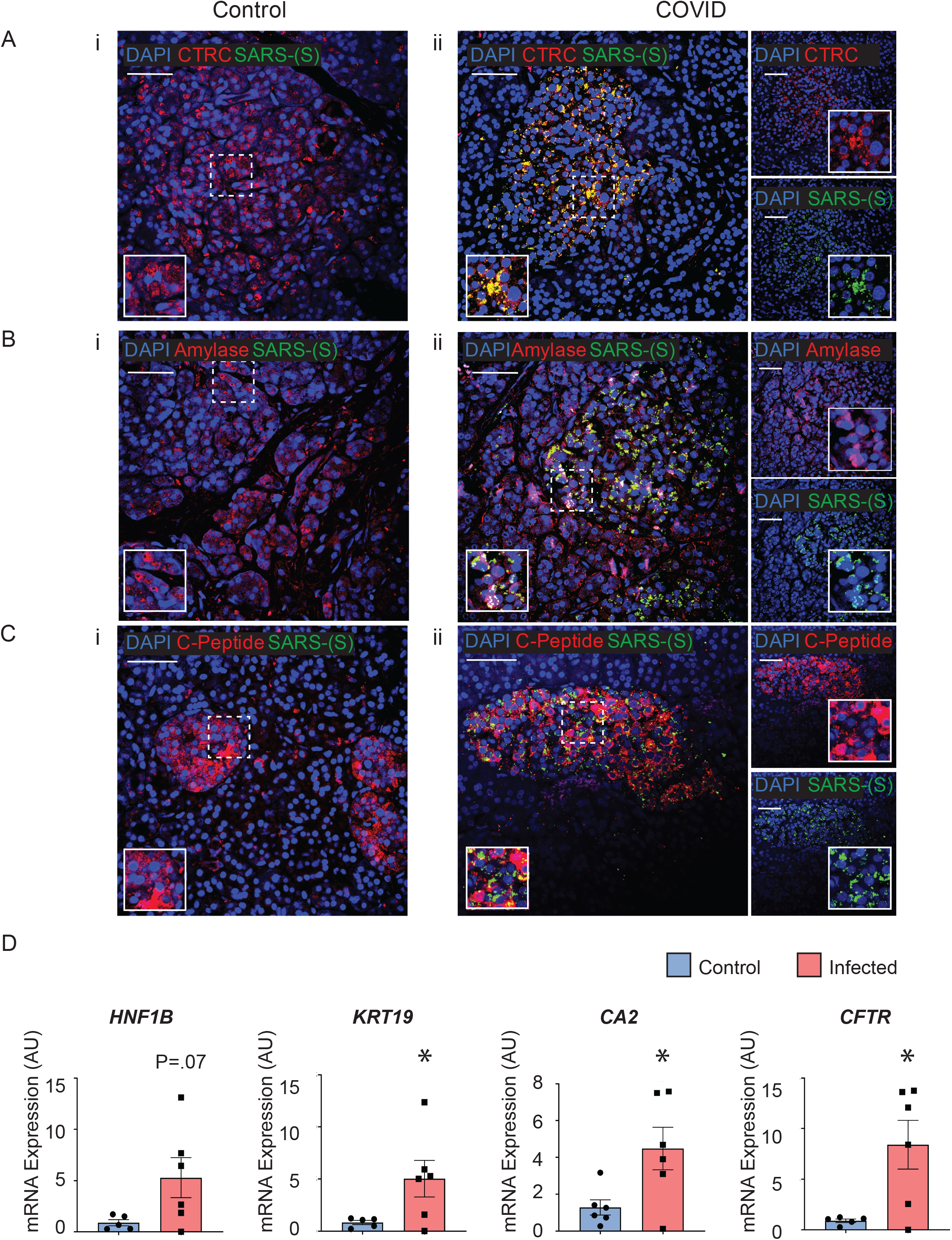
Post-mortem human pancreas shows SARS-CoV-2 infectivity and co-localization with multiple pancreatic cell types. Immunohistochemistry results from pancreatic tissues of patients with COVID-19 show SARS-CoV-2-Spike S1 staining in green and pancreatic acinar markers in red, such as **A ii**. Chymotrypsin, **B ii**. Amylase; and pancreatic endocrine marker **C ii**. C-peptide. **A-C i**. Corresponding sets of staining on non-COVID patients indicate no SARS-CoV-2-Spike S1 staining. **D**. Pancreatic tissues from COVID-19 patients (n=5) and Control subjects (n=6) were utilized for RNA extraction. Real-time qPCR results show increased mRNA expression of ductal markers *CA2, CFTR, KRT19* and *CFTR*. Data is shown as mean±SEM with statistical significance determined by unpaired two-tailed t-test. *p<0.05. Scale bar represents 50 µM.

To examine whether the infected pancreatic tissues from COVID-19 patients may have differential expression of specific genes, snap-frozen post-mortem tissues from COVID-19 patients and Control subjects were assessed changes in expression of pancreas-specific genes. Interestingly, mRNA expression of ductal markers was increased in the COVID-19 samples, such as *KRT19, CFTR, CA2*, and *HNF1B* as seen in Figure 6D. Other pancreatic and inflammatory markers were also tested and although showed trends of perturbations, they were not statistically different between the groups likely due to the higher variability in this subset of COVID-19 patients (**Suppl. Figure 6**). These results from COVID-19 and Control human tissues corroborate our novel results in iPan cell cultures, where we show that both pancreatic endocrine and exocrine cells can be infected by SARS-CoV-2, and the infection causes perturbations in pancreas-specific genes and the pancreatic cellular machinery.

## DISCUSSION

In this novel study, by observing gene and protein expression in live human iPSC-derived pancreatic cultures and post-mortem pancreatic tissue from COVID-19 patients in, we have described the ability of SARS-CoV-2 to infect pancreatic cells. Both approaches indicated that endocrine, acinar, and ductal cells within the pancreas are susceptible to SARS-CoV-2 infection. These cell types are known to express the ACE2 and TMPRSS2 transmembrane proteins recognized as the entry points for SARS-CoV-2 (Shang et al., 2020; Verdecchia et al., 2020). ACE2 and TMPRSS2 expression was observed as expected in the pancreatic cultures, thus providing an explanation for why pancreatic cells are permissive for SARS-CoV-2 infection. Recent clinical findings suggested pancreatic dysfunction following SARS-CoV-2 infection in COVID-19 patients (Apicella et al., 2020; Kusmartseva et al., 2020; Li et al., 2020; Liu et al., 2020). The iPSC-derived pancreatic cultures tested here were permissive for SARS-CoV-2 infection, as demonstrated by viral antigen expression and genomic replication. The cell culture observations were supported by similar results in post-mortem pancreatic tissues from COVID-19 patients. SARS-CoV-2 presence was detected clearly in the pancreas of infected patients, specifically co-localizing in exocrine ductal and acinar cells, as well as endocrine β-cells. Co-infection with β-cells containing islet clusters in post-mortem patients was notable given that diabetes and related complications are considered independent risk factors associated with higher mortality rates in COVID-19 (Holman et al., 2020).

Upon assessing cellular and molecular perturbations in iPSC-derived pancreatic cultures, it was observed that SARS-CoV-2 infected ductal cells in culture were enlarged and the nuclear compartmentalization of the ductal-specific transcription factor SOX9, which is typically observed in normal ductal cells, was absent. Instead, a cytoplasmic staining pattern, suggesting a mislocalization of SOX9 as a potential indicator of cellular stress imposed by the virus in infected cells (Chakravarty et al., 2011). The elongated morphology of the infected ductal cells is also a feature that is common in cancerous or inflamed epithelial cells converting to fibrotic cells undergoing a dynamic process known as epithelial-mesenchymal transition (EMT) (Kalluri and Weinberg, 2009; Krantz et al., 2012). Pancreatic tissues from COVID-19 patients showed perturbation with upregulation of key ductal genes such as *KRT19, CA2* and *CFTR*. Increased *KRT19* expression is associated with pancreas-specific intracellular stress in patients with poor prognosis of pancreatic ductal adenocarcinoma (PDAC) (Yao et al., 2017). Cytoskeletal protein responses are known to be dysregulated under conditions of intracellular stress imposed by a viral infection and similar disturbances have also been reported to be an important in both immune and adaptive immunity (Kopecki et al. 2016; Ong et al. 2020) Yet, further elucidation of the mechanisms of pancreatic injury involved in response to the SARS-CoV-2 infection in pancreatic ductal cells is necessary.

In infected acinar cell cultures, the proinflammatory genes such as *CXCL12* were upregulated. This gene, in particular, was found expressed ten-fold in blood plasma by (Xu et al., 2020), and is consistent with the overexpression of similar cytokines and chemokines widely seen during the cytokine storm frequently observed in COVID-19 patients (Coperchini et al., 2020; Song et al., 2020). Granular morphology was noted in all SARS-CoV-2 positive cells and could be representative of viral replication and transportation occurring in membrane vesicles. This would be consistent with previous findings that SARS-CoV-2 associates with the host endo membrane system, with 40% of SARS-CoV-2-interacting proteins having functions in the endomembrane system (Gordon et al., 2020).

Upon probing the pathways disrupted by SARS-CoV-2 upon infection of pancreatic cells, the most dramatic transcriptional change was the over-expression of transcriptional machinery and SRP-dependent protein-targeting processes. The transcripts contributing to these pathways were identified as hallmarks of viral replication by gene enrichment analysis. It has been previously reported that a common feature of coronaviruses is the use of virus-engineered double membrane vesicles from host cell components as a central site for viral RNA synthesis (Snijder et al., 2020; Wolff et al., 2020). The upregulation of SRP-protein targeting processes could be a reflection of host cell machinery being repurposed for viral replication. Conversely, the top three downregulated cellular components were nuclear. Other coronaviruses, including avian infectious bronchitis virus (IBV) and murine hepatitis virus (MHV), and SARS-CoV have been found to arrest cell cycle in the nucleus, leading to increased viral replication (Chen and Makino 2004; Dove et al. 2006; Yuan et al. 2006). Our results for the most upregulated and downregulated transcripts within the COVID-19-related Drug and Gene Set Library aligned with that of other coronaviruses, particularly SARS-CoV and MHV, with the top two downregulated transcripts being SARS-CoV-2 related. Taken all together, our results suggest that SARS-CoV-2, like other coronaviruses, elicits similar transcript-level signatures to promote viral replication in pancreatic cells. Future studies are necessary to confirm these mechanistic implications of these findings.

With this study, we provide novel patient-specific models for future mechanistic studies of SARS-CoV-2 impact on the pancreas This study supports the utility of iPSC-derived pancreatic cells as an excellent platform to explore the detrimental impacts of SARS-CoV-2 on pancreatic cells. Further, detailed studies are required to better understand and perhaps the 3D organoids and Organ-Chip systems can be utilized to explore the short-term and long-term detrimental effects SARS-CoV-2 has on the pancreas.

## Supporting information

Merged Supplemental Information

## Data Availability

All analysis and plotting scripts in R are available on github.com at github.com/Sareen-Lab/COVID.

## ACKNOWLEDGEMENTS AND FUNDING

We would like to acknowledge the support of the Cedars-Sinai Biomanufacturing Center facility and Cedars-Sinai Research Institute in supporting this project. This work was supported by Cedars-Sinai Programmatic Funds (DS). We would like to thank The David and Janet Polak Foundation. The funders had no role in study design, data collection and analysis, decision to publish, or preparation of the manuscript.

This research was also funded by UCLA DGSOM and Broad Stem Cell Research Center institutional award (OCRC #20-1) and (TRAN1COVID19-11975) to V.A. The following reagents were obtained through BEI Resources, NIAID, NIH: Polyclonal Anti-SARS Coronavirus (antiserum, Guinea Pig), NR-10361; The following reagent was deposited by the Centers for Disease Control and Prevention and obtained through BEI Resources, NIAID, NIH: SARS-Related Coronavirus 2, Isolate USA-WA1/2020, NR-52281. We are grateful to Barbara Dillon, UCLA High Containment Program Director for BSL3 work.

