## Supplementary material for "Deleterious effects of SARS-CoV-2 infection on human pancreatic cells": Merged Supplemental Information

† These authors contributed equally.

**This file includes:**

**Supplementary Figure and Table Legends.**

**Supplementary Figures 1-6.**

**Supplementary Tables** in a combined spreadsheet file containing the following supplementary tables (submitted as a separate .xlsx file).

#### SUPPLEMENTARY Figure AND Table LEGENDS

**Supplementary Figure 1. iPanEXO organoids exhibit ACE2 expression.** iPanEXO organoids exhibit polarization (E-cadherin), as well as markers of both pancreatic acinar (CTRC) and ductal cells (CK19, SOX9). These organoids exhibit ACE2 expression similar to adherent iPanEXO cultures. Scale bar represents 200 μm for large rectangle images, 50 μm for small square images, and 10 μm for zoomed panels adjacent to main images.

**Supplementary Figure 2. iPanEXO ductal cells express ACE2.** Day 3 post infected iPanEXO ductal cells exhibit ACE2 expression in SARS-CoV-2 infected and uninfected Mock control groups. The images below show the magnified regions of interest of DAPI and ACE2 staining in the dotted line boxes shown in the main panel for Mock and MOI 0.05 groups. Scale bar represents 200 μm.

**Supplementary Figure 3. C-peptide positive cells are infected by SARS-CoV-2.** iPanEXO acinar cultures contain small clusters of C-peptide protein expressing cells. A portion of these C-peptide positive cells were found to additionally express SARS-CoV-2-(S) on Day 3 (72 hours post-infection). Scale bar represents 130 μm, and 20 μm for zoomed panels adjacent to main images.

**Supplementary Figure 4. Inflammatory markers were not altered by SARS-CoV-2 infection at Day 1 and as well as IL1B and TNFA at Day 3**. **A.** RT-qPCR of infected and non-infected samples collected 24 hours (Day 1) after infection do not show significant change of mRNA expression of *CXCL12, IL1B, TNFA, NFKB1*, or *STAT3*. **B.** RT-qPCR of infected and non-infected samples collected 72 hours (Day 3) after infection do not show significant change of *IL1B* or *TNFA* mRNA expression. Data is shown as mean ± SEM with statistical significance determined by unpaired two-tailed t-test.

**Supplementary Figure 5. Post-Mortem Human Pancreas Staining of pancreatic ductal marker Cytokeratin 19.** Control and COVID-19 patients do not exhibit SARS-CoV-2 Spike S1 protein (green) in pancreatic ductal population, CK19 (red). Scale bar represents 50 μM.

**Supplementary Figure 6. mRNA expression of additional pancreatic and inflammatory markers from postmortem COVID-19 and Control patients.** Post-mortem pancreatic tissues from COVID-19 patients (n=6) and Control patients (n=6) were probed for mRNA expression of **A.**

endocrine markers *NGN3* and *INS*; **B.** acinar markers *CTRC, AMY1A, PTF1A and MIST1*; and **C.** inflammatory markers *IL1B, TNFA, CXCL12, NFKB1* and *STAT3*. Data is shown as mean ± SEM with statistical significance determined by unpaired two-tailed t-test.

**Supplementary Table 1.** Formulation of base media for culture of iPSC-derived iPan$^{EXO}$ Acinar and Ductal cells.

**Supplementary Table 2.** List of primer sequences used for Real-Time qPCR.

**Supplementary Table 3.** Expression of Viral Transcripts in RNAseq data.

**Supplementary Table 4.** Significant Differentially Expressed Genes Between SARS-CoV-2 Infected and Uninfected Cultures.

### Supplementary Figure 1

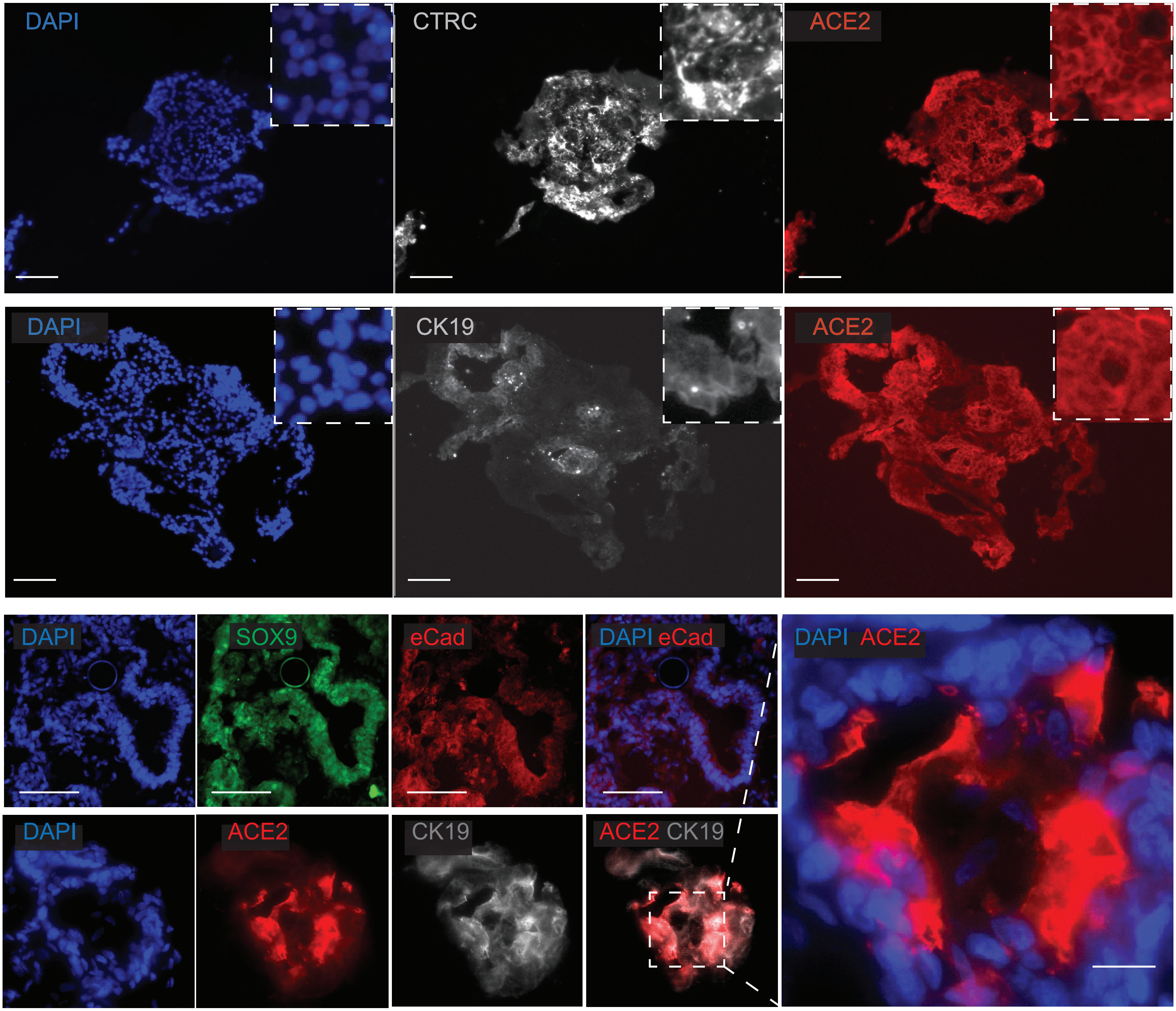


**Supplementary Fig 1. iPan$^{EXO}$ organoids exhibit ACE2 expression.** iPan$^{EXO}$ organoids exhibit polarization (E-cadherin), as well as markers of both pancreatic acinar (CTRC) and ductal cells (CK19, SOX9). These organoids exhibit ACE2 expression similar to adherent iPan$^{EXO}$ cultures. Scale bar represents 200 μm for large rectangle images, 50 μm for small square images, and 10 μm for zoomed panels adjacent to main images.

Supplementary Figure 2

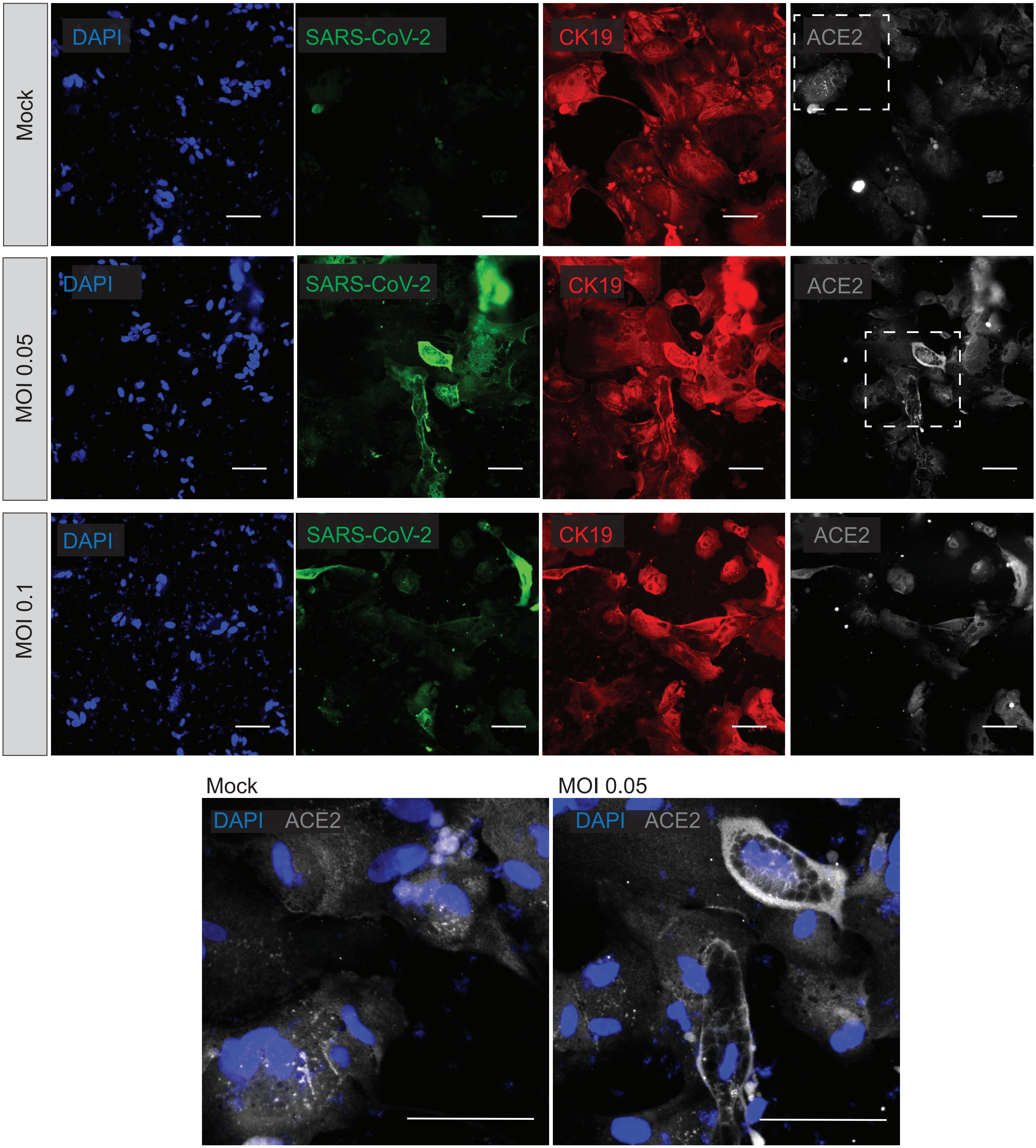


**Supplementary Fig 2. iPan$^{EXO}$ ductal cells express ACE2.** Day 3 post infected iPan$^{EXO}$ ductal cells exhibit ACE2 expression in SARS-CoV-2 infected and uninfected Mock control groups. The images below show the maginfied regions of interest of DAPI and ACE2 staining in the dotted line boxes shown in the main panel for Mock and MOI 0.05 groups. Scale bar represents 200 μm.

Supplementary Figure 3

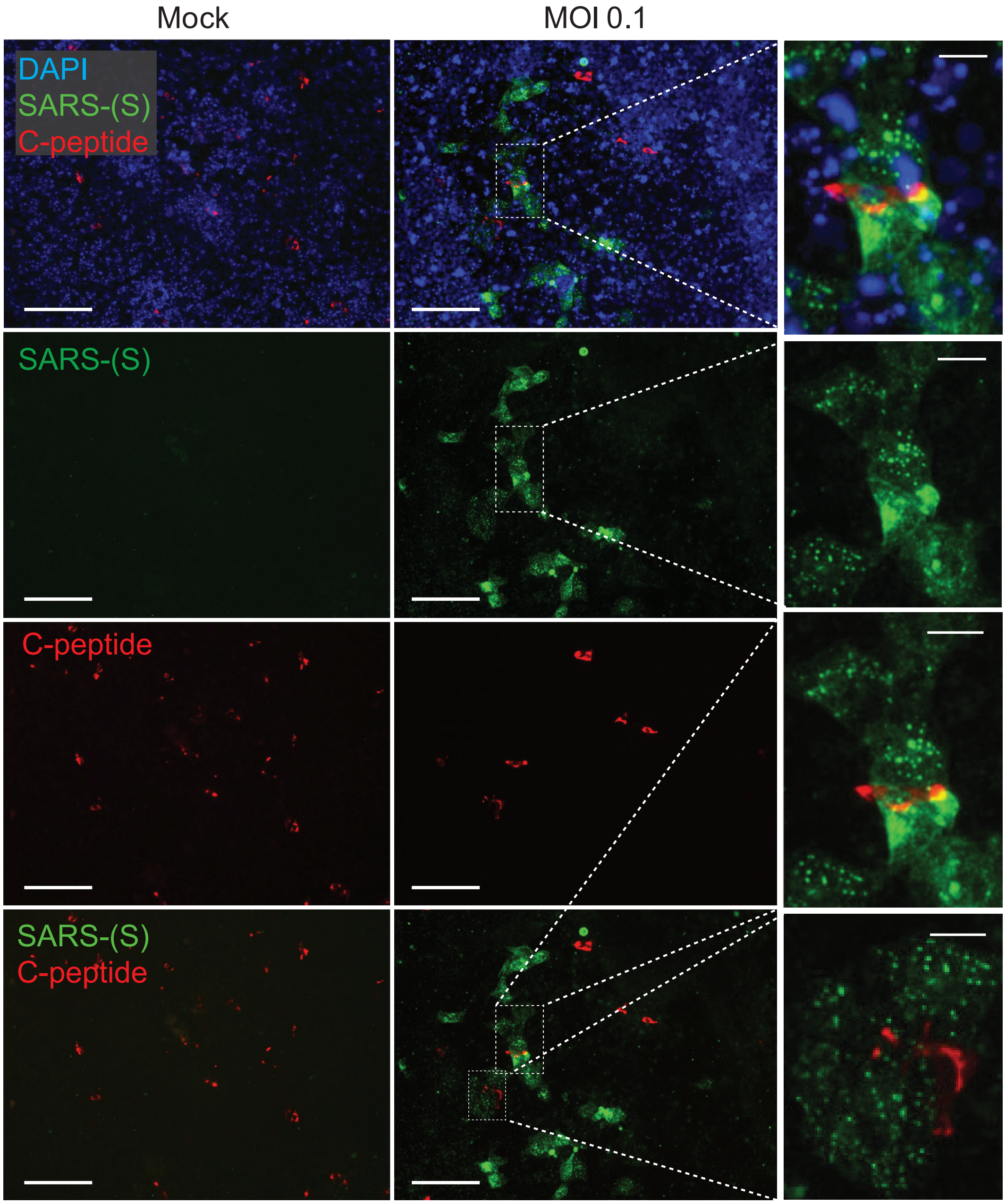


**Supplementary Fig 3. C-peptide positive cells are infected by SARS-CoV-2.** iPanEXO acinar cultures contain small clusters of C-peptide protein expressing cells. A portion of these C-peptide positive cells were found to additionally express SARS-(S) on Day 3 (72 hours post-infection). Scale bar represents 130 μm, and 20 μm for zoomed panels adjacent to main images.

Supplementary Figure 4

A

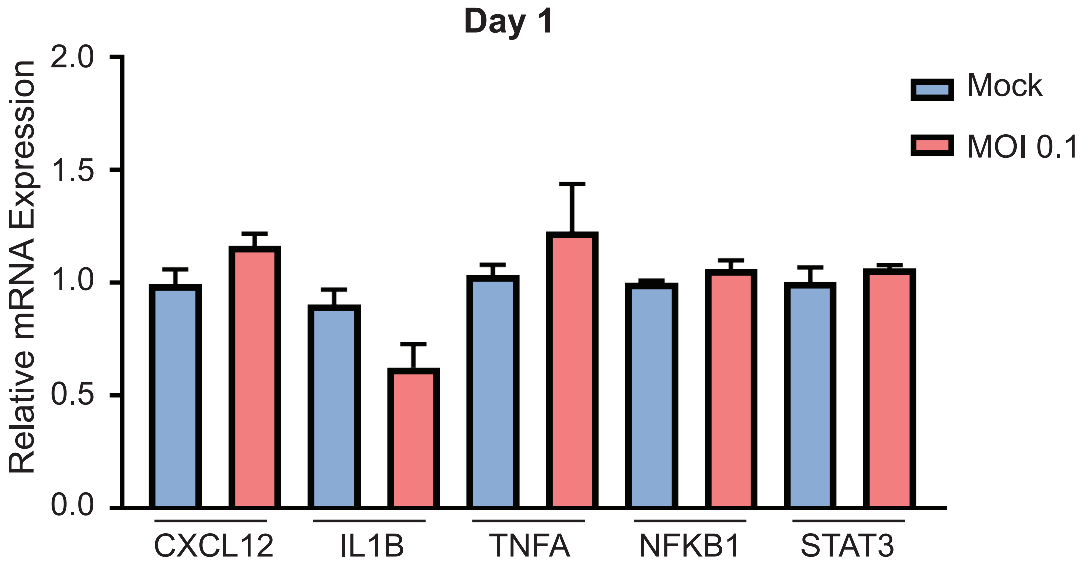


B

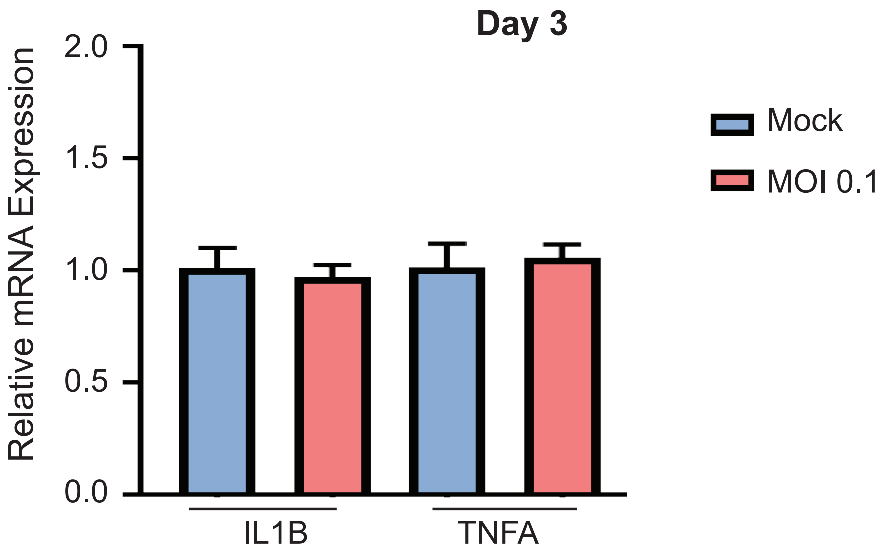


**Supplementary Fig 4. Inflammatory markers were not altered by SARS-CoV-2 infection at Day 1 and as well as *IL1B* and *TNFA* at Day 3. A.** RT-qPCR of infected and non-infected samples collected 24 hours (Day 1) after infection do not show significant change of mRNA expression of *CXCL12, IL1B, TNFA, NFKB1*, or *STAT3*. **B.** RT-qPCR of infected and non-infected samples collected 72 hours (Day 3) after infection do not show significant change of *IL1B* or *TNFA* mRNA expression. Data is shown as mean ± SEM with statistical significance determined by unpaired two-tailed t-test.

### Supplementary Figure 5

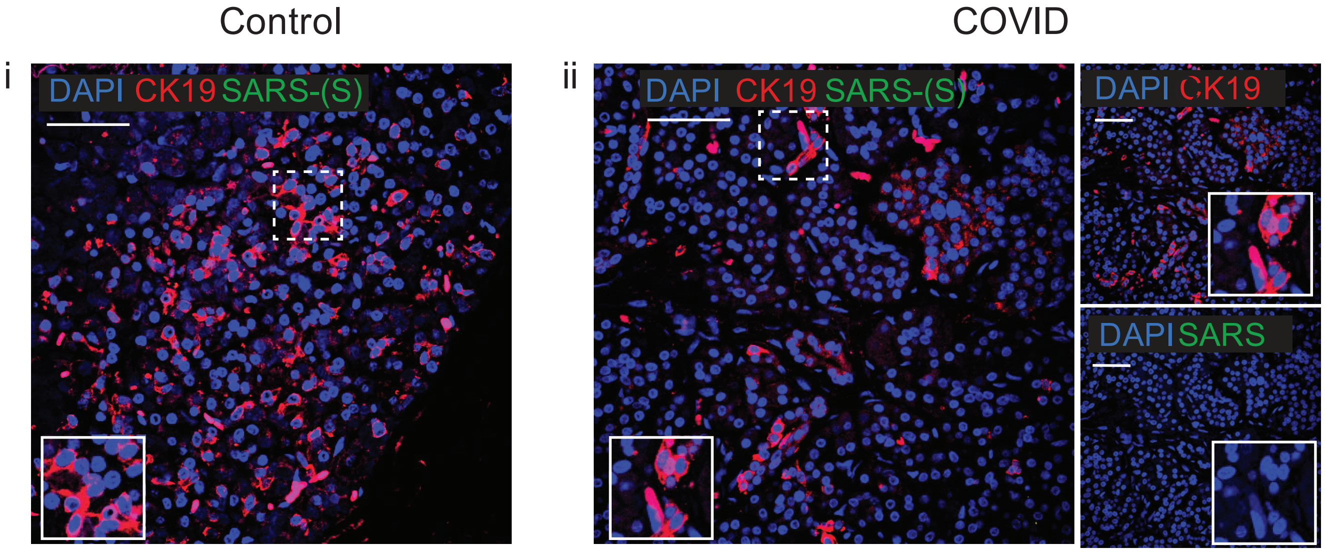


**Supplementary Fig 5. Post-Mortem Human Pancreas Staining of pancreatic ductal marker Cytokeratin 19.** Control and COVID-19 patients do not exhibit SARS-CoV-2 Spike S1 protein (green) in pancreatic ductal population, CK19 (red). Scale bar represents 50 μM.

Supplementary Figure 6

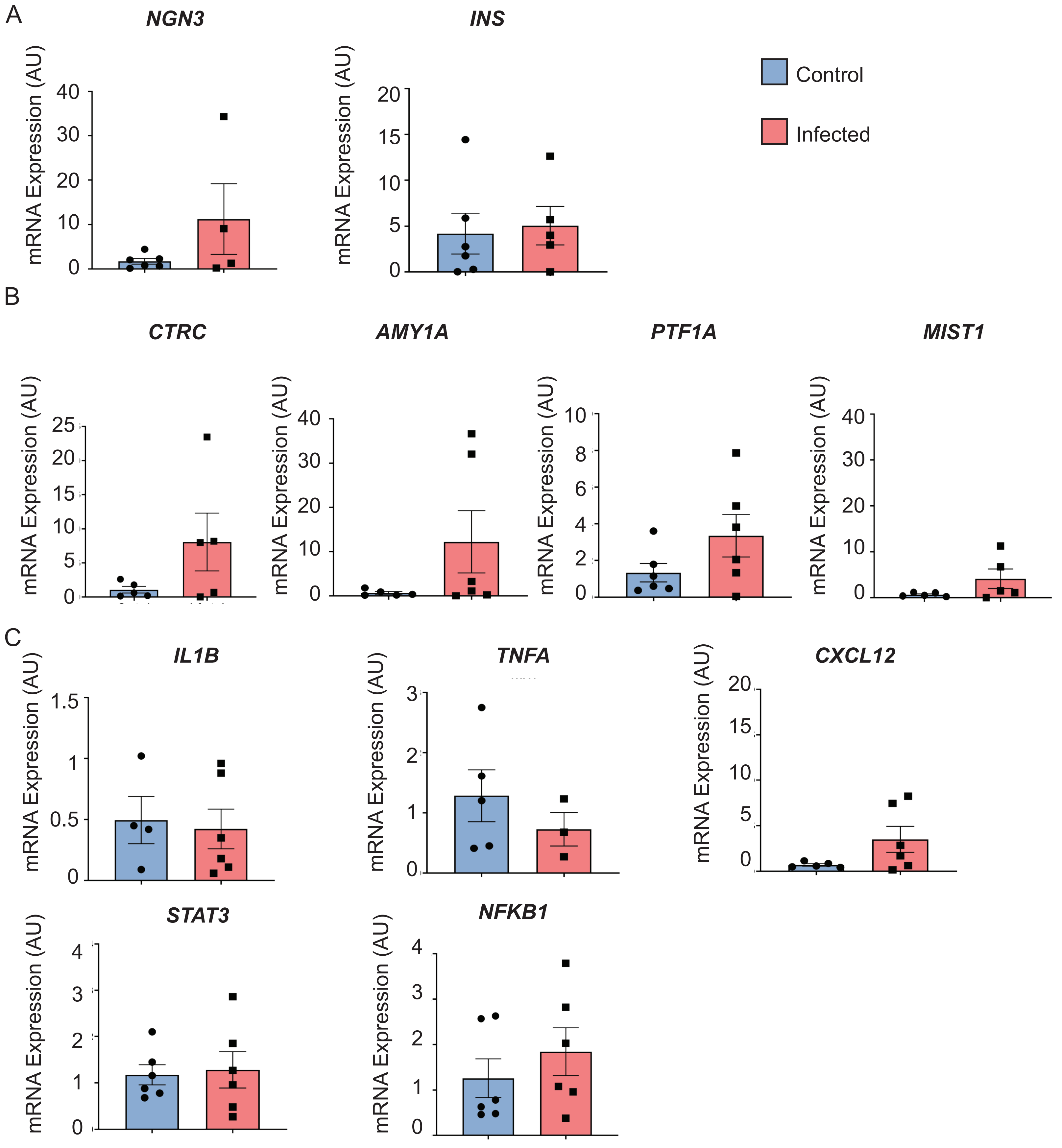


**Supplementary Fig 6. mRNA expression of additional pancreatic and inflammatory markers from post-mortem COVID-19 and Control patients.** Post-mortem pancreatic tissues from COVID-19 patients (n=6) and Control patients (n=6) were probed for mRNA expression of A. endocrine markers *NGN3* and *INS*; B. acinar markers *CTRC, AMY1A*, *PTF1A* and *MIST1*; and C. inflammatory markers *IL1B*, *TNFA*, *CXCL12*, *NFKB1* and *STAT3.* Data is shown as mean ± SEM with statistical significance determined by unpaired two-tailed t-test.

**Supplementary Table 1.** Formulation of base media for culture of iPSC-derived $iPan^{EXO}$ Acinar and Ductal cells.

| Phase I & II base medium | | |
|---|---|---|
| **Component** | **Amount** | **Final concentration** |
| MCDB131 (FisherScientific) | 98 ml | |
| Glutamax (FisherScientific) | 1 ml | 2 mM |
| Vitamin C (Sigma) | 100 µl | 250 µM |
| BSA (VWR) | 500 mg | 0.5% |
| $NaHCO_3$ (Sigma) | 150 mg | 1.5 g/L |
| Pen/Strep (Sigma) | 1 ml | 1% |
| **Phase III base medium** | | |
| **Component** | **Amount** | **Final concentration** |
| DMEM (ThermoFisher) | 98 ml | |
| Vitamin C | 100 µl | 250 µM |
| B27 supplement without Vitamin A (ThermoFisher) | 100 µl | 1% |
| Pen/Strep | 1 ml | 1% |

**Suppl. Table 2** List of primer sequences used for Real-Time qPCR.

| Gene | Forward (5´-3´) | Reverse (5´-3´) |
|---|---|---|
| *β-ACTIN* | TGACAGGATGCAGAAGGAGA | CGCTCAGGAGGAGCAATG |
| *GAPDH* | CCACCTTTGACGCTGGG | CATACCAGGAAATGAGCTTGACA |
| *RPL13* | GGCTAAACAGGTACTGCTGGG | AGGAAAGCCAGGTACTTCAACTT |
| *ACE2* | TCCAGACTCCGATCATCAAGC | TGCTCATGGTGTTCAGAATTGT |
| *TMPRSS2* | TATGAGAACCACGGGTATCAGT | CGTTGTAATCCTCGGAGCATACT |
| *SARS-CoV-2* | GACCCCAAAATCAGCGAAAT | TCTGGTTACTGCCAGTTGAATCTG |
| *NFKB1* | TCCATATTTGGGAAGGCCTGA | TTGAAGGTATGGGCCATCTGT |
| *CXCL12* | GACAAGTGTGCATTGACCCG | TCTCACATCTTGAACCTCTTGT |
| *IL1B* | AGCCATGGCAGAAGTACCTG | CCTGGAAGGAGCACTTCATCT |
| *TNFA* | CCCCAGGGACCTCTCTCTAAT | GCTTGAGGGTTTGCTACAAC |
| *STAT3* | ACCATTGACCTGCCGATGTC | AAGGTGAGGGACTCAAACTGC |
| *AMY2A* | TGTAACAATGTTGGGGTTCGT | CACTCACAGCGTTACCACAC |
| *CA2* | TCAGGACAAAGCAGTGCTCAA | TGTCCATCAAGTGAACCCCAG |
| *CFTR* | TGACCTTCTGCCTCTTACCA | CACTATCACTGGCACTGTTGC |
| *CTRC* | CCGCCTCTGGACCAACG | TGAAGCCCCACCAGTCAATC |
| *HNF1B* | GCACCCCTATGAAGACCCAG | GGACTGTCTGGTTGAATTGTCG |
| *INS* | AGGCCATCAAGCAGATCACT | TTCCCCGCACACTAGGTAGA |
| *KRT19* | GTTCACCAGCCGGACTGAA | GCAGGTCAGTAACCTCGGAC |
| *MIST1* | TCCAAGATCGAGACGCTCAC | TGCTGGACATGGTCAGGATG |
| *NGN3* | CGGTAGAAAGGATGACGCCT | GGTCACTTCGTCTTCCGAGG |
| *PTF1A* | TCGGGGCACCCGGTC | AGAGAGAGTGTCCTGCTAGGG |

**Supplementary Table 3.** Expression of Viral Transcripts in RNAseq data.

| | Day 1 Samples | | | | | | Day 3 Samples | | | | | |
|---|---|---|---|---|---|---|---|---|---|---|---|---|
| | Mock | | | Infected | | | Mock | | | Infected | | |
| | M1 | M2 | M3 | COV1 | COV2 | COV3 | M4 | M5 | M6 | COV4 | COV5 | COV6 |
| id_SARS_COV_2 _MT246667_ 1_1_261 | - | - | - | 162 | 192 | 198 | - | - | - | 616 | 823 | 815 |
| virus_E | - | - | - | 22 | 26 | 32 | - | - | - | 160 | 177 | 165 |
| virus_M | - | - | - | 265 | 276 | 293 | - | - | - | 1,488 | 1,828 | 1,891 |
| virus_N | - | 1 | - | 2,008 | 2,105 | 2,384 | - | - | - | 10,183 | 11,917 | 13,240 |
| virus_ORF10 | - | - | - | 84 | 72 | 65 | - | - | - | 422 | 485 | 434 |
| virus_ORF3a | - | - | - | 80 | 111 | 108 | - | - | - | 697 | 854 | 861 |
| virus_ORF6 | - | - | - | 25 | 29 | 31 | - | - | - | 148 | 167 | 136 |
| virus_ORF7a | - | - | - | 173 | 168 | 189 | - | - | - | 990 | 1,199 | 1,276 |
| virus_ORF8 | - | - | - | 193 | 209 | 251 | - | - | - | 1,244 | 1,531 | 1,479 |
| virus_S | - | - | - | 193 | 227 | 230 | - | - | - | 2,054 | 2,474 | 2,483 |
| virus_orf1ab | - | - | - | 325 | 336 | 333 | - | - | - | 2,333 | 2,837 | 3,176 |
| **Virus Total** | - | **1** | - | **3,530** | **3,751** | **4,114** | - | - | - | **20,335** | **24,292** | **25,956** |
| **Total Mapped Reads** | **2.95 E7** | **2.84 E7** | **3.26 E7** | **3.04 E7** | **3.18 E7** | **3.34 E7** | **2.89 E7** | **3.13 E7** | **2.46 E7** | **2.51 E7** | **3.02 E7** | **3.28 E7** |

Transcripts from the viral genome were detected within infected cell cultures. The count number of sequenced reads which mapped to the viral genome are listed above. M1, M2, M3, M4, M5, M6= indicate Mock conditions. COV1, COV2, COV3, COV4, COV5, COV6= indicate COVID-19 conditions.

**Supplementary Table 4.** Significant Differentially Expressed Genes Between SARS-CoV-2 Infected and Uninfected Cultures

| Ensembl ID | Gene | Description | Log2FC | Adjusted p-value |
|---|---|---|---|---|
| ENSG00000283765 | AC131160.1 | presenilin associated rhomboid like | 9.54 | 8.440E-12 |
| ENSG00000257921 | AC025165.3 | novel protein | 4.32 | 8.610E-06 |
| ENSG00000177984 | LCN15 | lipocalin 15 | 1.83 | 7.730E-05 |
| ENSG00000257390 | AC023055.1 | novel protein | 1.77 | 6.450E-05 |
| ENSG00000106483 | SFRP4 | secreted frizzled related protein 4 | 1.72 | 3.869E-04 |
| ENSG00000145721 | LIX1 | limb and CNS expressed 1 | 1.53 | 3.350E-06 |
| ENSG00000117318 | ID3 | inhibitor of DNA binding 3, HLH protein | 1.46 | 1.537E-02 |
| ENSG00000164107 | HAND2 | heart and neural crest derivatives expressed 2 | 1.44 | 3.270E-05 |
| ENSG00000101230 | ISM1 | isthmin 1 | 1.35 | 4.765E-02 |
| ENSG00000166482 | MFAP4 | microfibril associated protein 4 | 1.33 | 2.052E-02 |
| ENSG00000115263 | GCG | glucagon | 1.28 | 3.950E-07 |
| ENSG00000163359 | COL6A3 | collagen type VI alpha 3 chain | 1.24 | 5.520E-11 |
| ENSG00000124216 | SNAI1 | snail family transcriptional repressor 1 | 1.24 | 4.108E-02 |
| ENSG00000130700 | GATA5 | GATA binding protein 5 | 1.22 | 1.219E-03 |
| ENSG00000187634 | SAMD11 | sterile alpha motif domain containing 11 | 1.19 | 2.616E-03 |
| ENSG00000099960 | SLC7A4 | solute carrier family 7 member 4 | 1.14 | 5.305E-03 |
| ENSG00000287542 | AC098582.1 | HECT and RLD domain containing E3 ubiquitin protein ligase 3 | 1.14 | 1.097E-02 |
| ENSG00000079112 | CDH17 | cadherin 17 | 1.13 | 2.100E-06 |
| ENSG00000116132 | PRRX1 | paired related homeobox 1 | 1.13 | 1.578E-02 |
| ENSG00000159251 | ACTC1 | actin alpha cardiac muscle 1 | 1.12 | 2.330E-10 |
| ENSG00000090402 | SI | sucrase-isomaltase | 1.11 | 1.063E-02 |
| ENSG00000106366 | SERPINE1 | serpin family E member 1 | 1.11 | 2.575E-02 |
| ENSG00000169903 | TM4SF4 | transmembrane 4 L six family member 4 | 1.1 | 2.841E-04 |
| ENSG00000154188 | ANGPT1 | angiopoietin 1 | 1.1 | 8.086E-03 |
| ENSG00000240583 | AQP1 | aquaporin 1 (Colton blood group) | 1.1 | 3.698E-02 |
| ENSG00000254647 | INS | insulin | 1.09 | 7.400E-08 |
| ENSG00000179921 | GPBAR1 | G protein-coupled bile acid receptor 1 | 1.08 | 4.645E-02 |
| ENSG00000107731 | UNC5B | unc-5 netrin receptor B | 1.07 | 2.616E-03 |
| ENSG00000136698 | CFC1 | cripto, FRL-1, cryptic family 1 | 1.05 | 2.061E-03 |
| ENSG00000115738 | ID2 | inhibitor of DNA binding 2 | 1.02 | 1.590E-21 |
| ENSG00000151834 | GABRA2 | gamma-aminobutyric acid type A receptor subunit alpha2 | 1.02 | 7.830E-05 |
| ENSG00000120149 | MSX2 | msh homeobox 2 | 1 | 3.060E-06 |
| ENSG00000112175 | BMP5 | bone morphogenetic protein 5 | 1 | 2.690E-05 |
| ENSG00000157017 | GHRL | ghrelin and obestatin prepropeptide | 0.97 | 5.510E-21 |
| ENSG00000054598 | FOXC1 | forkhead box C1 | 0.96 | 1.959E-02 |

| | | | | |
|---|---|---|---|---|
| ENSG00000135111 | TBX3 | T-box transcription factor 3 | 0.95 | 1.458E-04 |
| ENSG00000213759 | UGT2B11 | UDP glucuronosyltransferase family 2 member B11 | 0.94 | 8.160E-06 |
| ENSG00000205038 | PKHD1L1 | PKHD1 like 1 | 0.91 | 2.843E-02 |
| ENSG00000172780 | RAB43 | RAB43, member RAS oncogene family | 0.87 | 6.920E-06 |
| ENSG00000248329 | APELA | apelin receptor early endogenous ligand | 0.87 | 2.530E-05 |
| ENSG00000183145 | RIPPLY3 | ripply transcriptional repressor 3 | 0.87 | 1.044E-02 |
| ENSG00000139515 | PDX1 | pancreatic and duodenal homeobox 1 | 0.84 | 1.632E-03 |
| ENSG00000165092 | ALDH1A1 | aldehyde dehydrogenase 1 family member A1 | 0.84 | 4.214E-02 |
| ENSG00000119946 | CNNM1 | cyclin and CBS domain divalent metal cation transport mediator 1 | 0.83 | 1.655E-02 |
| ENSG00000124875 | CXCL6 | C-X-C motif chemokine ligand 6 | 0.82 | 2.915E-02 |
| ENSG00000148344 | PTGES | prostaglandin E synthase | 0.8 | 5.740E-09 |
| ENSG00000159173 | TNNI1 | troponin I1, slow skeletal type | 0.8 | 4.764E-02 |
| ENSG00000163453 | IGFBP7 | insulin like growth factor binding protein 7 | 0.78 | 5.860E-10 |
| ENSG00000284395 | PERCC1 | proline and glutamate rich with coiled coil 1 | 0.75 | 3.322E-02 |
| ENSG00000172058 | SERF1A | small EDRK-rich factor 1A | 0.74 | 5.526E-04 |
| ENSG00000122691 | TWIST1 | twist family bHLH transcription factor 1 | 0.74 | 1.814E-02 |
| ENSG00000168542 | COL3A1 | collagen type III alpha 1 chain | 0.73 | 1.670E-08 |
| ENSG00000130829 | DUSP9 | dual specificity phosphatase 9 | 0.73 | 2.395E-02 |
| ENSG00000107562 | CXCL12 | C-X-C motif chemokine ligand 12 | 0.72 | 2.944E-02 |
| ENSG00000095739 | BAMBI | BMP and activin membrane bound inhibitor | 0.71 | 8.908E-04 |
| ENSG00000115363 | EVA1A | eva-1 homolog A, regulator of programmed cell death | 0.7 | 1.822E-02 |
| ENSG00000154162 | CDH12 | cadherin 12 | 0.69 | 1.607E-02 |
| ENSG00000136842 | TMOD1 | tropomodulin 1 | 0.68 | 3.036E-02 |
| ENSG00000101665 | SMAD7 | SMAD family member 7 | 0.67 | 9.751E-03 |
| ENSG00000103196 | CRISPLD2 | cysteine rich secretory protein LCCL domain containing 2 | 0.65 | 1.190E-05 |
| ENSG00000135744 | AGT | angiotensinogen | 0.65 | 3.273E-03 |
| ENSG00000129521 | EGLN3 | egl-9 family hypoxia inducible factor 3 | 0.64 | 7.398E-03 |
| ENSG00000130822 | PNCK | pregnancy up-regulated nonubiquitous CaM kinase | 0.64 | 1.737E-02 |
| ENSG00000181234 | TMEM132C | transmembrane protein 132C | 0.64 | 2.145E-02 |
| ENSG00000280987 | MATR3 | matrin 3 | 0.64 | 3.980E-02 |
| ENSG00000173826 | KCNH6 | potassium voltage-gated channel subfamily H member 6 | 0.64 | 4.975E-02 |
| ENSG00000135406 | PRPH | peripherin | 0.63 | 2.640E-02 |

| | | | | |
|---|---|---|---|---|
| ENSG00000185559 | DLK1 | delta like non-canonical Notch ligand 1 | 0.61 | 1.190E-05 |
| ENSG00000164237 | CMBL | carboxymethylenebutenolidase homolog | 0.61 | 8.304E-03 |
| ENSG00000124762 | CDKN1A | cyclin dependent kinase inhibitor 1A | 0.6 | 4.200E-06 |
| ENSG00000079689 | SCGN | secretagogin, EF-hand calcium binding protein | 0.6 | 2.048E-02 |
| ENSG00000187720 | THSD4 | thrombospondin type 1 domain containing 4 | 0.6 | 4.379E-02 |
| ENSG00000128606 | LRRC17 | leucine rich repeat containing 17 | 0.6 | 4.911E-02 |
| ENSG00000135480 | KRT7 | keratin 7 | 0.59 | 3.957E-03 |
| ENSG00000134013 | LOXL2 | lysyl oxidase like 2 | 0.59 | 2.567E-02 |
| ENSG00000115828 | QPCT | glutaminyl-peptide cyclotransferase | 0.59 | 4.308E-02 |
| ENSG00000265972 | TXNIP | thioredoxin interacting protein | 0.58 | 7.500E-11 |
| ENSG00000100604 | CHGA | chromogranin A | 0.58 | 8.629E-03 |
| ENSG00000122547 | EEPD1 | endonuclease/exonuclease/phosphatase family domain containing 1 | 0.58 | 3.800E-02 |
| ENSG00000146674 | IGFBP3 | insulin like growth factor binding protein 3 | 0.56 | 1.483E-02 |
| ENSG00000090013 | BLVRB | biliverdin reductase B | 0.56 | 2.145E-02 |
| ENSG00000175426 | PCSK1 | proprotein convertase subtilisin/kexin type 1 | 0.56 | 4.549E-02 |
| ENSG00000001626 | CFTR | CF transmembrane conductance regulator | 0.55 | 1.403E-02 |
| ENSG00000103260 | METRN | meteorin, glial cell differentiation regulator | 0.55 | 3.572E-02 |
| ENSG00000166165 | CKB | creatine kinase B | 0.53 | 1.560E-10 |
| ENSG00000130429 | ARPC1B | actin related protein 2/3 complex subunit 1B | 0.53 | 6.667E-04 |
| ENSG00000108821 | COL1A1 | collagen type I alpha 1 chain | 0.53 | 2.778E-02 |
| ENSG00000115221 | ITGB6 | integrin subunit beta 6 | 0.52 | 3.816E-04 |
| ENSG00000187840 | EIF4EBP1 | eukaryotic translation initiation factor 4E binding protein 1 | 0.52 | 3.645E-03 |
| ENSG00000105143 | SLC1A6 | solute carrier family 1 member 6 | 0.52 | 1.011E-02 |
| ENSG00000140450 | ARRDC4 | arrestin domain containing 4 | 0.51 | 3.950E-07 |
| ENSG00000154734 | ADAMTS1 | ADAM metallopeptidase with thrombospondin type 1 motif 1 | 0.51 | 2.841E-04 |
| ENSG00000178057 | NDUFAF3 | NADH:ubiquinone oxidoreductase complex assembly factor 3 | 0.51 | 5.676E-03 |
| ENSG00000133027 | PEMT | phosphatidylethanolamine N-methyltransferase | 0.51 | 8.706E-03 |
| ENSG00000065057 | NTHL1 | nth like DNA glycosylase 1 | 0.51 | 2.785E-02 |
| ENSG00000169220 | RGS14 | regulator of G protein signaling 14 | 0.51 | 4.238E-02 |
| ENSG00000049130 | KITLG | KIT ligand | 0.5 | 9.760E-05 |
| ENSG00000123358 | NR4A1 | nuclear receptor subfamily 4 group A member 1 | 0.5 | 4.394E-03 |
| ENSG00000165066 | NKX6-3 | NK6 homeobox 3 | 0.5 | 7.698E-03 |

| | | | | |
|---|---|---|---|---|
| ENSG00000110104 | CCDC86 | coiled-coil domain containing 86 | 0.5 | 1.454E-02 |
| ENSG00000183048 | SLC25A10 | solute carrier family 25 member 10 | 0.5 | 3.839E-02 |
| ENSG00000175793 | SFN | stratifin | 0.5 | 4.660E-02 |
| ENSG00000157005 | SST | somatostatin | 0.49 | 2.263E-02 |
| ENSG00000063241 | ISOC2 | isochorismatase domain containing 2 | 0.49 | 2.372E-02 |
| ENSG00000126218 | F10 | coagulation factor X | 0.49 | 4.306E-02 |
| ENSG00000112576 | CCND3 | cyclin D3 | 0.48 | 1.097E-02 |
| ENSG00000165272 | AQP3 | aquaporin 3 (Gill blood group) | 0.48 | 1.233E-02 |
| ENSG00000112276 | BVES | blood vessel epicardial substance | 0.48 | 3.575E-02 |
| ENSG00000171159 | C9orf16 | chromosome 9 open reading frame 16 | 0.47 | 1.302E-02 |
| ENSG00000134258 | VTCN1 | V-set domain containing T cell activation inhibitor 1 | 0.46 | 9.290E-07 |
| ENSG00000205544 | TMEM256 | transmembrane protein 256 | 0.46 | 4.130E-06 |
| ENSG00000162878 | PKDCC | protein kinase domain containing, cytoplasmic | 0.46 | 1.600E-05 |
| ENSG00000151090 | THRB | thyroid hormone receptor beta | 0.46 | 4.802E-03 |
| ENSG00000120708 | TGFBI | transforming growth factor beta induced | 0.46 | 5.522E-03 |
| ENSG00000152082 | MZT2B | mitotic spindle organizing protein 2B | 0.46 | 1.195E-02 |
| ENSG00000183010 | PYCR1 | pyrroline-5-carboxylate reductase 1 | 0.46 | 2.313E-02 |
| ENSG00000196975 | ANXA4 | annexin A4 | 0.45 | 2.776E-03 |
| ENSG00000122863 | CHST3 | carbohydrate sulfotransferase 3 | 0.45 | 1.328E-02 |
| ENSG00000127561 | SYNGR3 | synaptogyrin 3 | 0.45 | 4.771E-02 |
| ENSG00000161958 | FGF11 | fibroblast growth factor 11 | 0.44 | 1.728E-04 |
| ENSG00000087245 | MMP2 | matrix metallopeptidase 2 | 0.44 | 1.674E-02 |
| ENSG00000141526 | SLC16A3 | solute carrier family 16 member 3 | 0.44 | 4.737E-02 |
| ENSG00000151726 | ACSL1 | acyl-CoA synthetase long chain family member 1 | 0.43 | 6.520E-08 |
| ENSG00000083845 | RPS5 | ribosomal protein S5 | 0.43 | 3.050E-07 |
| ENSG00000141448 | GATA6 | GATA binding protein 6 | 0.43 | 1.020E-03 |
| ENSG00000168000 | BSCL2 | BSCL2 lipid droplet biogenesis associated, seipin | 0.43 | 4.453E-03 |
| ENSG00000109861 | CTSC | cathepsin C | 0.42 | 1.632E-03 |
| ENSG00000181026 | AEN | apoptosis enhancing nuclease | 0.42 | 2.537E-03 |
| ENSG00000034152 | MAP2K3 | mitogen-activated protein kinase kinase 3 | 0.42 | 6.236E-03 |
| ENSG00000101220 | C20orf27 | chromosome 20 open reading frame 27 | 0.42 | 1.086E-02 |
| ENSG00000125170 | DOK4 | docking protein 4 | 0.42 | 1.737E-02 |
| ENSG00000172731 | LRRC20 | leucine rich repeat containing 20 | 0.41 | 1.972E-02 |
| ENSG00000120885 | CLU | clusterin | 0.41 | 4.179E-02 |
| ENSG00000124145 | SDC4 | syndecan 4 | 0.4 | 1.821E-04 |
| ENSG00000180914 | OXTR | oxytocin receptor | 0.4 | 3.074E-04 |
| ENSG00000181788 | SIAH2 | siah E3 ubiquitin protein ligase 2 | 0.4 | 1.172E-02 |
| ENSG00000117691 | NENF | neudesin neurotrophic factor | 0.4 | 1.271E-02 |

| | | | | |
|---|---|---|---|---|
| ENSG00000155254 | MARVELD1 | MARVEL domain containing 1 | 0.4 | 1.420E-02 |
| ENSG00000115884 | SDC1 | syndecan 1 | 0.39 | 4.190E-05 |
| ENSG00000131469 | RPL27 | ribosomal protein L27 | 0.39 | 1.458E-04 |
| ENSG00000267855 | NDUFA7 | NADH:ubiquinone oxidoreductase subunit A7 | 0.39 | 7.333E-03 |
| ENSG00000105372 | RPS19 | ribosomal protein S19 | 0.38 | 1.979E-04 |
| ENSG00000074370 | ATP2A3 | ATPase sarcoplasmic/endoplasmic reticulum Ca2+ transporting 3 | 0.38 | 7.087E-04 |
| ENSG00000176946 | THAP4 | THAP domain containing 4 | 0.38 | 1.777E-03 |
| ENSG00000145685 | LHFPL2 | LHFPL tetraspan subfamily member 2 | 0.38 | 1.985E-02 |
| ENSG00000140937 | CDH11 | cadherin 11 | 0.37 | 4.134E-03 |
| ENSG00000110108 | TMEM109 | transmembrane protein 109 | 0.37 | 4.823E-03 |
| ENSG00000165716 | DIPK1B | divergent protein kinase domain 1B | 0.37 | 1.097E-02 |
| ENSG00000011260 | UTP18 | UTP18 small subunit processome component | 0.37 | 1.362E-02 |
| ENSG00000142327 | RNPEPL1 | arginyl aminopeptidase like 1 | 0.37 | 3.783E-02 |
| ENSG00000140564 | FURIN | furin, paired basic amino acid cleaving enzyme | 0.37 | 4.496E-02 |
| ENSG00000164587 | RPS14 | ribosomal protein S14 | 0.36 | 4.090E-06 |
| ENSG00000054148 | PHPT1 | phosphohistidine phosphatase 1 | 0.36 | 1.455E-04 |
| ENSG00000189143 | CLDN4 | claudin 4 | 0.36 | 2.943E-04 |
| ENSG00000090776 | EFNB1 | ephrin B1 | 0.36 | 5.803E-04 |
| ENSG00000088756 | ARHGAP28 | Rho GTPase activating protein 28 | 0.36 | 1.097E-02 |
| ENSG00000130332 | LSM7 | LSM7 homolog, U6 small nuclear RNA and mRNA degradation associated | 0.36 | 1.134E-02 |
| ENSG00000183172 | SMDT1 | single-pass membrane protein with aspartate rich tail 1 | 0.36 | 2.001E-02 |
| ENSG00000095906 | NUBP2 | nucleotide binding protein 2 | 0.36 | 2.111E-02 |
| ENSG00000166595 | CIAO2B | cytosolic iron-sulfur assembly component 2B | 0.36 | 2.700E-02 |
| ENSG00000275993 | SIK1B | salt inducible kinase 1B (putative) | 0.36 | 4.496E-02 |
| ENSG00000105640 | RPL18A | ribosomal protein L18a | 0.35 | 1.930E-05 |
| ENSG00000103257 | SLC7A5 | solute carrier family 7 member 5 | 0.35 | 3.070E-05 |
| ENSG00000164638 | SLC29A4 | solute carrier family 29 member 4 | 0.35 | 4.031E-03 |
| ENSG00000148834 | GSTO1 | glutathione S-transferase omega 1 | 0.35 | 1.619E-02 |
| ENSG00000177556 | ATOX1 | antioxidant 1 copper chaperone | 0.35 | 2.472E-02 |
| ENSG00000131043 | AAR2 | AAR2 splicing factor | 0.35 | 3.839E-02 |
| ENSG00000077238 | IL4R | interleukin 4 receptor | 0.35 | 4.911E-02 |
| ENSG00000240972 | MIF | macrophage migration inhibitory factor | 0.34 | 1.720E-03 |
| ENSG00000138449 | SLC40A1 | solute carrier family 40 member 1 | 0.34 | 2.488E-03 |
| ENSG00000233927 | RPS28 | ribosomal protein S28 | 0.34 | 3.645E-03 |
| ENSG00000165672 | PRDX3 | peroxiredoxin 3 | 0.34 | 1.030E-02 |
| ENSG00000112559 | MDFI | MyoD family inhibitor | 0.34 | 1.441E-02 |
| ENSG00000006282 | SPATA20 | spermatogenesis associated 20 | 0.34 | 2.012E-02 |

| | | | | |
|---|---|---|---|---|
| ENSG00000214063 | TSPAN4 | tetraspanin 4 | 0.34 | 3.790E-02 |
| ENSG00000123179 | EBPL | EBP like | 0.34 | 4.310E-02 |
| ENSG00000121900 | TMEM54 | transmembrane protein 54 | 0.34 | 4.423E-02 |
| ENSG00000109475 | RPL34 | ribosomal protein L34 | 0.33 | 5.175E-04 |
| ENSG00000068001 | HYAL2 | hyaluronidase 2 | 0.33 | 6.018E-04 |
| ENSG00000198931 | APRT | adenine phosphoribosyltransferase | 0.33 | 1.097E-02 |
| ENSG00000221983 | UBA52 | ubiquitin A-52 residue ribosomal protein fusion product 1 | 0.33 | 1.221E-02 |
| ENSG00000109736 | MFSD10 | major facilitator superfamily domain containing 10 | 0.33 | 1.354E-02 |
| ENSG00000185664 | PMEL | premelanosome protein | 0.33 | 1.483E-02 |
| ENSG00000120256 | LRP11 | LDL receptor related protein 11 | 0.33 | 2.376E-02 |
| ENSG00000105193 | RPS16 | ribosomal protein S16 | 0.33 | 2.966E-02 |
| ENSG00000171604 | CXXC5 | CXXC finger protein 5 | 0.33 | 3.389E-02 |
| ENSG00000123992 | DNPEP | aspartyl aminopeptidase | 0.33 | 3.786E-02 |
| ENSG00000152078 | TLCD4 | TLC domain containing 4 | 0.33 | 3.839E-02 |
| ENSG00000063177 | RPL18 | ribosomal protein L18 | 0.33 | 4.310E-02 |
| ENSG00000167695 | TLCD3A | TLC domain containing 3A | 0.33 | 4.516E-02 |
| ENSG00000239264 | TXNDC5 | thioredoxin domain containing 5 | 0.32 | 5.180E-05 |
| ENSG00000178202 | POGLUT3 | protein O-glucosyltransferase 3 | 0.32 | 3.686E-03 |
| ENSG00000147403 | RPL10 | ribosomal protein L10 | 0.32 | 4.353E-03 |
| ENSG00000204392 | LSM2 | LSM2 homolog, U6 small nuclear RNA and mRNA degradation associated | 0.32 | 6.337E-03 |
| ENSG00000130635 | COL5A1 | collagen type V alpha 1 chain | 0.32 | 1.029E-02 |
| ENSG00000115268 | RPS15 | ribosomal protein S15 | 0.32 | 1.043E-02 |
| ENSG00000125995 | ROMO1 | reactive oxygen species modulator 1 | 0.32 | 2.587E-02 |
| ENSG00000137801 | THBS1 | thrombospondin 1 | 0.32 | 4.529E-02 |
| ENSG00000105677 | TMEM147 | transmembrane protein 147 | 0.32 | 4.549E-02 |
| ENSG00000133243 | BTBD2 | BTB domain containing 2 | 0.32 | 4.600E-02 |
| ENSG00000205542 | TMSB4X | thymosin beta 4 X-linked | 0.31 | 1.260E-05 |
| ENSG00000188986 | NELFB | negative elongation factor complex member B | 0.31 | 2.907E-03 |
| ENSG00000166886 | NAB2 | NGFI-A binding protein 2 | 0.31 | 6.148E-03 |
| ENSG00000143811 | PYCR2 | pyrroline-5-carboxylate reductase 2 | 0.31 | 1.097E-02 |
| ENSG00000106153 | CHCHD2 | coiled-coil-helix-coiled-coil-helix domain containing 2 | 0.31 | 1.497E-02 |
| ENSG00000071082 | RPL31 | ribosomal protein L31 | 0.31 | 1.498E-02 |
| ENSG00000102265 | TIMP1 | TIMP metallopeptidase inhibitor 1 | 0.31 | 4.109E-02 |
| ENSG00000198840 | MT-ND3 | mitochondrially encoded NADH:ubiquinone oxidoreductase core subunit 3 | 0.31 | 4.472E-02 |
| ENSG00000198242 | RPL23A | ribosomal protein L23a | 0.3 | 3.005E-04 |
| ENSG00000116857 | TMEM9 | transmembrane protein 9 | 0.3 | 7.041E-04 |
| ENSG00000165283 | STOML2 | stomatin like 2 | 0.3 | 2.877E-03 |
| ENSG00000100814 | CCNB1IP1 | cyclin B1 interacting protein 1 | 0.3 | 3.112E-03 |
| ENSG00000198918 | RPL39 | ribosomal protein L39 | 0.3 | 3.645E-03 |

| | | | | |
|---|---|---|---|---|
| ENSG00000143543 | JTB | jumping translocation breakpoint | 0.3 | 8.694E-03 |
| ENSG00000204388 | HSPA1B | heat shock protein family A (Hsp70) member 1B | 0.3 | 2.133E-02 |
| ENSG00000149806 | FAU | FAU ubiquitin like and ribosomal protein S30 fusion | 0.3 | 4.050E-02 |
| ENSG00000117419 | ERI3 | ERI1 exoribonuclease family member 3 | 0.3 | 4.975E-02 |
| ENSG00000127184 | COX7C | cytochrome c oxidase subunit 7C | 0.29 | 1.539E-04 |
| ENSG00000101444 | AHCY | adenosylhomocysteinase | 0.29 | 3.576E-04 |
| ENSG00000132507 | EIF5A | eukaryotic translation initiation factor 5A | 0.29 | 3.643E-04 |
| ENSG00000149948 | HMGA2 | high mobility group AT-hook 2 | 0.29 | 4.938E-04 |
| ENSG00000164292 | RHOBTB3 | Rho related BTB domain containing 3 | 0.29 | 3.871E-03 |
| ENSG00000136240 | KDELR2 | KDEL endoplasmic reticulum protein retention receptor 2 | 0.29 | 6.834E-03 |
| ENSG00000231500 | RPS18 | ribosomal protein S18 | 0.29 | 8.305E-03 |
| ENSG00000084207 | GSTP1 | glutathione S-transferase pi 1 | 0.29 | 1.100E-02 |
| ENSG00000177830 | CHID1 | chitinase domain containing 1 | 0.29 | 1.362E-02 |
| ENSG00000183098 | GPC6 | glypican 6 | 0.29 | 1.407E-02 |
| ENSG00000171858 | RPS21 | ribosomal protein S21 | 0.29 | 1.656E-02 |
| ENSG00000170892 | TSEN34 | tRNA splicing endonuclease subunit 34 | 0.29 | 2.785E-02 |
| ENSG00000162244 | RPL29 | ribosomal protein L29 | 0.29 | 2.902E-02 |
| ENSG00000172809 | RPL38 | ribosomal protein L38 | 0.29 | 3.449E-02 |
| ENSG00000198951 | NAGA | alpha-N-acetylgalactosaminidase | 0.29 | 3.843E-02 |
| ENSG00000167543 | TP53I13 | tumor protein p53 inducible protein 13 | 0.29 | 4.549E-02 |
| ENSG00000176101 | SSNA1 | SS nuclear autoantigen 1 | 0.29 | 4.660E-02 |
| ENSG00000198763 | MT-ND2 | mitochondrially encoded NADH:ubiquinone oxidoreductase core subunit 2 | 0.28 | 4.149E-03 |
| ENSG00000184840 | TMED9 | transmembrane p24 trafficking protein 9 | 0.28 | 6.480E-03 |
| ENSG00000117448 | AKR1A1 | aldo-keto reductase family 1 member A1 | 0.28 | 8.696E-03 |
| ENSG00000172354 | GNB2 | G protein subunit beta 2 | 0.28 | 9.782E-03 |
| ENSG00000170889 | RPS9 | ribosomal protein S9 | 0.28 | 1.254E-02 |
| ENSG00000099797 | TECR | trans-2,3-enoyl-CoA reductase | 0.28 | 1.607E-02 |
| ENSG00000134291 | TMEM106C | transmembrane protein 106C | 0.28 | 2.109E-02 |
| ENSG00000179091 | CYC1 | cytochrome c1 | 0.28 | 3.246E-02 |
| ENSG00000168003 | SLC3A2 | solute carrier family 3 member 2 | 0.28 | 3.914E-02 |
| ENSG00000126768 | TIMM17B | translocase of inner mitochondrial membrane 17B | 0.28 | 4.011E-02 |
| ENSG00000164692 | COL1A2 | collagen type I alpha 2 chain | 0.28 | 4.459E-02 |
| ENSG00000137288 | UQCC2 | ubiquinol-cytochrome c reductase complex assembly factor 2 | 0.28 | 4.879E-02 |

| | | | | |
|---|---|---|---|---|
| ENSG00000100316 | RPL3 | ribosomal protein L3 | 0.27 | 5.324E-04 |
| ENSG00000143761 | ARF1 | ADP ribosylation factor 1 | 0.27 | 1.410E-02 |
| ENSG00000167291 | TBC1D16 | TBC1 domain family member 16 | 0.27 | 2.108E-02 |
| ENSG00000101182 | PSMA7 | proteasome 20S subunit alpha 7 | 0.27 | 3.337E-02 |
| ENSG00000171551 | ECEL1 | endothelin converting enzyme like 1 | 0.27 | 4.448E-02 |
| ENSG00000247596 | TWF2 | twinfilin actin binding protein 2 | 0.27 | 4.517E-02 |
| ENSG00000107438 | PDLIM1 | PDZ and LIM domain 1 | 0.26 | 4.823E-03 |
| ENSG00000175390 | EIF3F | eukaryotic translation initiation factor 3 subunit F | 0.26 | 6.513E-03 |
| ENSG00000013306 | SLC25A39 | solute carrier family 25 member 39 | 0.26 | 1.254E-02 |
| ENSG00000125166 | GOT2 | glutamic-oxaloacetic transaminase 2 | 0.26 | 1.853E-02 |
| ENSG00000243678 | NME2 | NME/NM23 nucleoside diphosphate kinase 2 | 0.26 | 1.862E-02 |
| ENSG00000172992 | DCAKD | dephospho-CoA kinase domain containing | 0.26 | 2.450E-02 |
| ENSG00000204628 | RACK1 | receptor for activated C kinase 1 | 0.26 | 2.533E-02 |
| ENSG00000124614 | RPS10 | ribosomal protein S10 | 0.26 | 4.052E-02 |
| ENSG00000198755 | RPL10A | ribosomal protein L10a | 0.25 | 2.062E-03 |
| ENSG00000075618 | FSCN1 | fascin actin-bundling protein 1 | 0.25 | 8.756E-03 |
| ENSG00000213741 | RPS29 | ribosomal protein S29 | 0.25 | 1.362E-02 |
| ENSG00000110651 | CD81 | CD81 molecule | 0.25 | 2.376E-02 |
| ENSG00000112514 | CUTA | cutA divalent cation tolerance homolog | 0.25 | 2.657E-02 |
| ENSG00000182899 | RPL35A | ribosomal protein L35a | 0.25 | 2.902E-02 |
| ENSG00000167283 | ATP5MG | ATP synthase membrane subunit g | 0.25 | 3.823E-02 |
| ENSG00000126267 | COX6B1 | cytochrome c oxidase subunit 6B1 | 0.25 | 4.004E-02 |
| ENSG00000140931 | CMTM3 | CKLF like MARVEL transmembrane domain containing 3 | 0.25 | 4.599E-02 |
| ENSG00000116685 | KIAA2013 | KIAA2013 | 0.25 | 4.737E-02 |
| ENSG00000084623 | EIF3I | eukaryotic translation initiation factor 3 subunit I | 0.24 | 2.575E-02 |
| ENSG00000049239 | H6PD | hexose-6-phosphate dehydrogenase/glucose 1-dehydrogenase | 0.24 | 2.727E-02 |
| ENSG00000149923 | PPP4C | protein phosphatase 4 catalytic subunit | 0.24 | 3.574E-02 |
| ENSG00000169230 | PRELID1 | PRELI domain containing 1 | 0.24 | 3.644E-02 |
| ENSG00000104635 | SLC39A14 | solute carrier family 39 member 14 | 0.24 | 3.812E-02 |
| ENSG00000110987 | BCL7A | BAF chromatin remodeling complex subunit BCL7A | 0.24 | 3.839E-02 |
| ENSG00000160691 | SHC1 | SHC adaptor protein 1 | 0.24 | 4.341E-02 |
| ENSG00000125817 | CENPB | centromere protein B | 0.24 | 4.455E-02 |
| ENSG00000131165 | CHMP1A | charged multivesicular body protein 1A | 0.24 | 4.459E-02 |
| ENSG00000168028 | RPSA | ribosomal protein SA | 0.23 | 8.189E-04 |
| ENSG00000089157 | RPLP0 | ribosomal protein lateral stalk subunit P0 | 0.23 | 9.471E-04 |

| | | | | |
|---|---|---|---|---|
| ENSG00000075624 | ACTB | actin beta | 0.23 | 1.078E-03 |
| ENSG00000117592 | PRDX6 | peroxiredoxin 6 | 0.23 | 6.148E-03 |
| ENSG00000137409 | MTCH1 | mitochondrial carrier 1 | 0.23 | 1.674E-02 |
| ENSG00000155380 | SLC16A1 | solute carrier family 16 member 1 | 0.23 | 1.674E-02 |
| ENSG00000124172 | ATP5F1E | ATP synthase F1 subunit epsilon | 0.23 | 1.845E-02 |
| ENSG00000125835 | SNRPB | small nuclear ribonucleoprotein polypeptides B and B1 | 0.23 | 2.850E-02 |
| ENSG00000130726 | TRIM28 | tripartite motif containing 28 | 0.23 | 3.356E-02 |
| ENSG00000100138 | SNU13 | small nuclear ribonucleoprotein 13 | 0.23 | 4.333E-02 |
| ENSG00000130638 | ATXN10 | ataxin 10 | 0.22 | 2.450E-02 |
| ENSG00000104738 | MCM4 | minichromosome maintenance complex component 4 | 0.22 | 2.510E-02 |
| ENSG00000143418 | CERS2 | ceramide synthase 2 | 0.22 | 3.839E-02 |
| ENSG00000197324 | LRP10 | LDL receptor related protein 10 | 0.22 | 4.023E-02 |
| ENSG00000184009 | ACTG1 | actin gamma 1 | 0.22 | 4.139E-02 |
| ENSG00000100129 | EIF3L | eukaryotic translation initiation factor 3 subunit L | 0.21 | 9.743E-03 |
| ENSG00000173726 | TOMM20 | translocase of outer mitochondrial membrane 20 | 0.21 | 2.403E-02 |
| ENSG00000110700 | RPS13 | ribosomal protein S13 | 0.21 | 3.511E-02 |
| ENSG00000073111 | MCM2 | minichromosome maintenance complex component 2 | 0.21 | 3.773E-02 |
| ENSG00000105202 | FBL | fibrillarin | 0.21 | 4.496E-02 |
| ENSG00000168268 | NT5DC2 | 5'-nucleotidase domain containing 2 | 0.21 | 4.597E-02 |
| ENSG00000139644 | TMBIM6 | transmembrane BAX inhibitor motif containing 6 | 0.2 | 1.370E-02 |
| ENSG00000108298 | RPL19 | ribosomal protein L19 | 0.2 | 3.836E-02 |
| ENSG00000184117 | NIPSNAP1 | nipsnap homolog 1 | 0.19 | 2.275E-02 |
| ENSG00000115484 | CCT4 | chaperonin containing TCP1 subunit 4 | 0.19 | 4.548E-02 |
| ENSG00000075415 | SLC25A3 | solute carrier family 25 member 3 | 0.19 | 4.764E-02 |
| ENSG00000112306 | RPS12 | ribosomal protein S12 | 0.18 | 4.307E-02 |
| ENSG00000198938 | MT-CO3 | mitochondrially encoded cytochrome c oxidase III | 0.16 | 4.939E-02 |
| ENSG00000277277 | FAM243B | family with sequence similarity 243 member B | -4.64 | 9.782E-03 |
| ENSG00000272741 | AC069257.4 | novel protein | -3.89 | 2.523E-02 |
| ENSG00000285258 | ATXN7 | NA | -2.84 | 1.410E-02 |
| ENSG00000187642 | PERM1 | PPARGC1 and ESRR induced regulator, muscle 1 | -2.83 | 2.327E-02 |
| ENSG00000185040 | SPDYE16 | speedy/RINGO cell cycle regulator family member E16 | -2.81 | 2.893E-02 |
| ENSG00000261732 | AL031708.1 | novel protein | -2.27 | 3.960E-07 |
| ENSG00000226763 | SRRM5 | serine/arginine repetitive matrix 5 | -2.2 | 3.183E-02 |

| | | | | |
|---|---|---|---|---|
| ENSG00000249773 | AC092647.5 | novel zinc finger protein 713 (ZNF713) and mitochondrial ribosomal protein S17 (MRPS17) protein | -2.08 | 2.882E-04 |
| ENSG00000205238 | SPDYE2 | speedy/RINGO cell cycle regulator family member E2 | -2.04 | 6.480E-03 |
| ENSG00000127364 | TAS2R4 | taste 2 receptor member 4 | -1.97 | 2.133E-04 |
| ENSG00000215018 | COL28A1 | collagen type XXVIII alpha 1 chain | -1.94 | 9.782E-03 |
| ENSG00000272916 | AC022400.7 | novel transcript | -1.89 | 8.340E-06 |
| ENSG00000089692 | LAG3 | lymphocyte activating 3 | -1.83 | 3.585E-02 |
| ENSG00000100078 | PLA2G3 | phospholipase A2 group III | -1.7 | 4.423E-02 |
| ENSG00000170092 | SPDYE5 | speedy/RINGO cell cycle regulator family member E5 | -1.69 | 7.398E-03 |
| ENSG00000285130 | AL358113.1 | novel protein | -1.65 | 2.583E-02 |
| ENSG00000186399 | GOLGA8R | golgin A8 family member R | -1.42 | 4.496E-02 |
| ENSG00000170160 | CCDC144A | coiled-coil domain containing 144A | -1.37 | 6.796E-03 |
| ENSG00000286053 | ASDURF | ASNSD1 upstream reading frame | -1.37 | 3.350E-03 |
| ENSG00000172005 | MAL | mal, T cell differentiation protein | -1.36 | 3.356E-02 |
| ENSG00000240053 | LY6G5B | lymphocyte antigen 6 family member G5B | -1.34 | 9.330E-06 |
| ENSG00000164707 | SLC13A4 | solute carrier family 13 member 4 | -1.27 | 5.084E-03 |
| ENSG00000164796 | CSMD3 | CUB and Sushi multiple domains 3 | -1.26 | 5.433E-03 |
| ENSG00000145113 | MUC4 | mucin 4, cell surface associated | -1.21 | 1.675E-03 |
| ENSG00000242028 | HYPK | huntingtin interacting protein K | -1.17 | 1.250E-05 |
| ENSG00000204889 | KRT40 | keratin 40 | -1.16 | 1.461E-02 |
| ENSG00000170370 | EMX2 | empty spiracles homeobox 2 | -1.14 | 6.610E-05 |
| ENSG00000215252 | GOLGA8B | golgin A8 family member B | -1.14 | 1.780E-06 |
| ENSG00000166762 | CATSPER2 | cation channel sperm associated 2 | -1.13 | 2.065E-04 |
| ENSG00000135976 | ANKRD36 | ankyrin repeat domain 36 | -1.12 | 1.410E-13 |
| ENSG00000166432 | ZMAT1 | zinc finger matrin-type 1 | -1.11 | 4.970E-05 |
| ENSG00000274272 | AC069281.2 | novel transcript | -1.09 | 4.945E-02 |
| ENSG00000250067 | YJEFN3 | YjeF N-terminal domain containing 3 | -1.07 | 8.400E-06 |
| ENSG00000145832 | SLC25A48 | solute carrier family 25 member 48 | -1.06 | 5.218E-04 |
| ENSG00000118997 | DNAH7 | dynein axonemal heavy chain 7 | -1.05 | 1.032E-03 |
| ENSG00000066923 | STAG3 | stromal antigen 3 | -1.02 | 5.430E-09 |
| ENSG00000213462 | ERV3-1 | endogenous retrovirus group 3 member 1, envelope | -1.02 | 5.190E-12 |
| ENSG00000163126 | ANKRD23 | ankyrin repeat domain 23 | -1.01 | 1.012E-02 |
| ENSG00000009694 | TENM1 | teneurin transmembrane protein 1 | -1 | 3.873E-03 |
| ENSG00000118515 | SGK1 | serum/glucocorticoid regulated kinase 1 | -0.99 | 4.003E-02 |
| ENSG00000285053 | TBCE | tubulin folding cofactor E | -0.99 | 2.930E-02 |
| ENSG00000230551 | AC021078.1 | novel transcript | -0.98 | 6.540E-07 |
| ENSG00000184206 | GOLGA6L4 | golgin A6 family like 4 | -0.97 | 1.101E-03 |
| ENSG00000262152 | LINC00514 | long intergenic non-protein coding RNA 514 | -0.96 | 3.682E-02 |
| ENSG00000123572 | NRK | Nik related kinase | -0.96 | 1.822E-02 |

| | | | | |
|---|---|---|---|---|
| ENSG00000084674 | APOB | apolipoprotein B | -0.95 | 1.550E-05 |
| ENSG00000139537 | CCDC65 | coiled-coil domain containing 65 | -0.94 | 4.764E-02 |
| ENSG00000243708 | PLA2G4B | phospholipase A2 group IVB | -0.94 | 4.376E-02 |
| ENSG00000274808 | TBC1D3B | TBC1 domain family member 3B | -0.94 | 4.315E-02 |
| ENSG00000243696 | AC006254.1 | novel MUSTN1-ITIH4 readthrough | -0.94 | 4.741E-03 |
| ENSG00000082196 | C1QTNF3 | C1q and TNF related 3 | -0.93 | 3.840E-02 |
| ENSG00000185864 | NPIPB4 | nuclear pore complex interacting protein family member B4 | -0.93 | 3.730E-05 |
| ENSG00000273154 | AL121845.3 | novel protein, ZGPAT-LIME1 readthrough | -0.92 | 1.613E-02 |
| ENSG00000174501 | ANKRD36C | ankyrin repeat domain 36C | -0.92 | 8.990E-05 |
| ENSG00000181722 | ZBTB20 | zinc finger and BTB domain containing 20 | -0.91 | 5.320E-06 |
| ENSG00000213903 | LTB4R | leukotriene B4 receptor | -0.89 | 3.267E-04 |
| ENSG00000186166 | CCDC84 | coiled-coil domain containing 84 | -0.89 | 1.926E-04 |
| ENSG00000246922 | UBAP1L | ubiquitin associated protein 1 like | -0.85 | 1.607E-02 |
| ENSG00000114270 | COL7A1 | collagen type VII alpha 1 chain | -0.84 | 9.330E-06 |
| ENSG00000139190 | VAMP1 | vesicle associated membrane protein 1 | -0.83 | 4.285E-02 |
| ENSG00000101104 | PABPC1L | poly(A) binding protein cytoplasmic 1 like | -0.82 | 4.790E-10 |
| ENSG00000114857 | NKTR | natural killer cell triggering receptor | -0.82 | 7.110E-17 |
| ENSG00000254995 | STX16-NPEPL | STX16-NPEPL1 readthrough (NMD candidate) | -0.81 | 7.399E-03 |
| ENSG00000173559 | NABP1 | nucleic acid binding protein 1 | -0.8 | 4.802E-03 |
| ENSG00000241489 | AC244197.3 | novel protein | -0.79 | 2.570E-08 |
| ENSG00000175265 | GOLGA8A | golgin A8 family member A | -0.78 | 1.700E-21 |
| ENSG00000171811 | CFAP46 | cilia and flagella associated protein 46 | -0.77 | 4.338E-02 |
| ENSG00000198556 | ZNF789 | zinc finger protein 789 | -0.76 | 1.498E-02 |
| ENSG00000156042 | CFAP70 | cilia and flagella associated protein 70 | -0.75 | 2.785E-02 |
| ENSG00000169246 | NPIPB3 | nuclear pore complex interacting protein family member B3 | -0.75 | 5.040E-10 |
| ENSG00000204248 | COL11A2 | collagen type XI alpha 2 chain | -0.74 | 4.441E-02 |
| ENSG00000197748 | CFAP43 | cilia and flagella associated protein 43 | -0.74 | 3.905E-02 |
| ENSG00000157765 | SLC34A2 | solute carrier family 34 member 2 | -0.74 | 3.210E-02 |
| ENSG00000160201 | U2AF1 | U2 small nuclear RNA auxiliary factor 1 | -0.74 | 2.847E-02 |
| ENSG00000278662 | GOLGA6L10 | golgin A6 family like 10 | -0.74 | 2.098E-03 |
| ENSG00000188738 | FSIP2 | fibrous sheath interacting protein 2 | -0.74 | 1.958E-03 |
| ENSG00000204172 | AGAP9 | ArfGAP with GTPase domain, ankyrin repeat and PH domain 9 | -0.73 | 1.166E-02 |
| ENSG00000206530 | CFAP44 | cilia and flagella associated protein 44 | -0.72 | 2.485E-02 |
| ENSG00000111664 | GNB3 | G protein subunit beta 3 | -0.72 | 2.388E-02 |

| | | | | |
|---|---|---|---|---|
| ENSG00000135905 | DOCK10 | dedicator of cytokinesis 10 | -0.7 | 2.069E-03 |
| ENSG00000180626 | ZNF594 | zinc finger protein 594 | -0.7 | 6.325E-04 |
| ENSG00000062370 | ZNF112 | zinc finger protein 112 | -0.69 | 4.198E-02 |
| ENSG00000214029 | ZNF891 | zinc finger protein 891 | -0.68 | 7.991E-04 |
| ENSG00000166532 | RIMKLB | ribosomal modification protein rimK like family member B | -0.68 | 2.570E-07 |
| ENSG00000177990 | DPY19L2 | dpy-19 like 2 | -0.67 | 4.304E-02 |
| ENSG00000120327 | PCDHB14 | protocadherin beta 14 | -0.67 | 3.487E-02 |
| ENSG00000104894 | CD37 | CD37 molecule | -0.67 | 3.259E-02 |
| ENSG00000185829 | ARL17A | ADP ribosylation factor like GTPase 17A | -0.67 | 1.691E-02 |
| ENSG00000182310 | SPACA6 | sperm acrosome associated 6 | -0.67 | 4.149E-03 |
| ENSG00000152926 | ZNF117 | zinc finger protein 117 | -0.67 | 4.890E-08 |
| ENSG00000128000 | ZNF780B | zinc finger protein 780B | -0.66 | 3.376E-02 |
| ENSG00000139631 | CSAD | cysteine sulfinic acid decarboxylase | -0.66 | 3.869E-04 |
| ENSG00000167615 | LENG8 | leukocyte receptor cluster member 8 | -0.65 | 5.803E-04 |
| ENSG00000163945 | UVSSA | UV stimulated scaffold protein A | -0.65 | 2.430E-07 |
| ENSG00000184619 | KRBA2 | KRAB-A domain containing 2 | -0.64 | 2.575E-02 |
| ENSG00000166436 | TRIM66 | tripartite motif containing 66 | -0.64 | 2.060E-02 |
| ENSG00000214021 | TTLL3 | tubulin tyrosine ligase like 3 | -0.64 | 1.212E-02 |
| ENSG00000145824 | CXCL14 | C-X-C motif chemokine ligand 14 | -0.64 | 2.301E-04 |
| ENSG00000275740 | AC091959.3 | novel readthrough transcript | -0.63 | 2.843E-02 |
| ENSG00000172345 | STARD5 | StAR related lipid transfer domain containing 5 | -0.62 | 1.872E-02 |
| ENSG00000185513 | L3MBTL1 | L3MBTL histone methyl-lysine binding protein 1 | -0.62 | 7.850E-07 |
| ENSG00000204410 | MSH5 | mutS homolog 5 | -0.6 | 2.388E-02 |
| ENSG00000137474 | MYO7A | myosin VIIA | -0.6 | 3.612E-03 |
| ENSG00000186472 | PCLO | piccolo presynaptic cytomatrix protein | -0.6 | 2.109E-03 |
| ENSG00000167524 | RSKR | ribosomal protein S6 kinase related | -0.6 | 6.001E-04 |
| ENSG00000189367 | KIAA0408 | KIAA0408 | -0.59 | 4.014E-02 |
| ENSG00000196912 | ANKRD36B | ankyrin repeat domain 36B | -0.58 | 4.044E-02 |
| ENSG00000157423 | HYDIN | HYDIN axonemal central pair apparatus protein | -0.58 | 6.143E-03 |
| ENSG00000241404 | EGFL8 | EGF like domain multiple 8 | -0.58 | 5.224E-03 |
| ENSG00000130518 | IQCN | IQ motif containing N | -0.57 | 4.409E-02 |
| ENSG00000265590 | C21orf59-TCP | CFAP298-TCP10L readthrough | -0.57 | 8.744E-03 |
| ENSG00000104899 | AMH | anti-Mullerian hormone | -0.56 | 1.959E-02 |
| ENSG00000162825 | NBPF20 | NBPF member 20 | -0.56 | 1.690E-02 |
| ENSG00000162601 | MYSM1 | Myb like, SWIRM and MPN domains 1 | -0.56 | 3.645E-03 |
| ENSG00000127914 | AKAP9 | A-kinase anchoring protein 9 | -0.56 | 1.821E-04 |
| ENSG00000197978 | GOLGA6L9 | golgin A6 family like 9 | -0.56 | 9.290E-07 |
| ENSG00000243716 | NPIPB5 | nuclear pore complex interacting protein family member B5 | -0.56 | 5.190E-12 |

| | | | | |
|---|---|---|---|---|
| ENSG00000154265 | ABCA5 | ATP binding cassette subfamily A member 5 | -0.54 | 1.003E-02 |
| ENSG00000121454 | LHX4 | LIM homeobox 4 | -0.54 | 6.480E-03 |
| ENSG00000156313 | RPGR | retinitis pigmentosa GTPase regulator | -0.54 | 3.209E-03 |
| ENSG00000197774 | EME2 | essential meiotic structure-specific endonuclease subunit 2 | -0.54 | 2.133E-04 |
| ENSG00000008311 | AASS | aminoadipate-semialdehyde synthase | -0.53 | 4.338E-02 |
| ENSG00000145476 | CYP4V2 | cytochrome P450 family 4 subfamily V member 2 | -0.53 | 3.183E-02 |
| ENSG00000137496 | IL18BP | interleukin 18 binding protein | -0.53 | 1.256E-02 |
| ENSG00000214193 | SH3D21 | SH3 domain containing 21 | -0.53 | 3.635E-03 |
| ENSG00000188234 | AGAP4 | ArfGAP with GTPase domain, ankyrin repeat and PH domain 4 | -0.53 | 5.324E-04 |
| ENSG00000163728 | TTC14 | tetratricopeptide repeat domain 14 | -0.53 | 1.728E-04 |
| ENSG00000126775 | ATG14 | autophagy related 14 | -0.52 | 4.088E-03 |
| ENSG00000054654 | SYNE2 | spectrin repeat containing nuclear envelope protein 2 | -0.52 | 1.020E-03 |
| ENSG00000116560 | SFPQ | splicing factor proline and glutamine rich | -0.52 | 4.740E-06 |
| ENSG00000137393 | RNF144B | ring finger protein 144B | -0.51 | 4.696E-02 |
| ENSG00000138658 | ZGRF1 | zinc finger GRF-type containing 1 | -0.51 | 4.355E-02 |
| ENSG00000080823 | MOK | MOK protein kinase | -0.51 | 2.282E-02 |
| ENSG00000059588 | TARBP1 | TAR (HIV-1) RNA binding protein 1 | -0.51 | 1.865E-03 |
| ENSG00000176155 | CCDC57 | coiled-coil domain containing 57 | -0.51 | 2.522E-04 |
| ENSG00000215788 | TNFRSF25 | TNF receptor superfamily member 25 | -0.5 | 4.179E-02 |
| ENSG00000166801 | FAM111A | family with sequence similarity 111 member A | -0.5 | 2.584E-03 |
| ENSG00000144749 | LRIG1 | leucine rich repeats and immunoglobulin like domains 1 | -0.5 | 1.819E-03 |
| ENSG00000055609 | KMT2C | lysine methyltransferase 2C | -0.5 | 1.549E-04 |
| ENSG00000198625 | MDM4 | MDM4 regulator of p53 | -0.5 | 8.710E-05 |
| ENSG00000138002 | IFT172 | intraflagellar transport 172 | -0.49 | 3.576E-04 |
| ENSG00000189042 | ZNF567 | zinc finger protein 567 | -0.48 | 4.229E-02 |
| ENSG00000232593 | KANTR | KDM5C adjacent transcript | -0.48 | 1.073E-02 |
| ENSG00000196876 | SCN8A | sodium voltage-gated channel alpha subunit 8 | -0.48 | 8.696E-03 |
| ENSG00000121310 | ECHDC2 | enoyl-CoA hydratase domain containing 2 | -0.48 | 4.149E-03 |
| ENSG00000163660 | CCNL1 | cyclin L1 | -0.48 | 1.600E-05 |
| ENSG00000102908 | NFAT5 | nuclear factor of activated T cells 5 | -0.48 | 3.980E-07 |
| ENSG00000256591 | AP003108.2 | novel transcript | -0.47 | 4.691E-02 |
| ENSG00000128849 | CGNL1 | cingulin like 1 | -0.47 | 4.517E-02 |
| ENSG00000160298 | C21orf58 | chromosome 21 open reading frame 58 | -0.47 | 2.450E-02 |

| | | | | |
|---|---|---|---|---|
| ENSG00000008196 | TFAP2B | transcription factor AP-2 beta | -0.47 | 2.001E-02 |
| ENSG00000113916 | BCL6 | BCL6 transcription repressor | -0.47 | 1.233E-02 |
| ENSG00000148143 | ZNF462 | zinc finger protein 462 | -0.47 | 1.169E-02 |
| ENSG00000233024 | NPIPA9 | nuclear pore complex interacting protein family, member A9 | -0.47 | 5.997E-03 |
| ENSG00000092094 | OSGEP | O-sialoglycoprotein endopeptidase | -0.47 | 2.882E-04 |
| ENSG00000108848 | LUC7L3 | LUC7 like 3 pre-mRNA splicing factor | -0.47 | 1.930E-07 |
| ENSG00000224078 | SNHG14 | small nucleolar RNA host gene 14 | -0.47 | 1.030E-07 |
| ENSG00000131711 | MAP1B | microtubule associated protein 1B | -0.47 | 4.570E-09 |
| ENSG00000151914 | DST | dystonin | -0.47 | 9.020E-11 |
| ENSG00000184205 | TSPYL2 | TSPY like 2 | -0.46 | 1.811E-02 |
| ENSG00000273136 | NBPF26 | NBPF member 26 | -0.46 | 8.527E-03 |
| ENSG00000144026 | ZNF514 | zinc finger protein 514 | -0.46 | 4.031E-03 |
| ENSG00000175455 | CCDC14 | coiled-coil domain containing 14 | -0.46 | 6.050E-07 |
| ENSG00000165323 | FAT3 | FAT atypical cadherin 3 | -0.45 | 2.927E-02 |
| ENSG00000165359 | INTS6L | integrator complex subunit 6 like | -0.45 | 2.785E-02 |
| ENSG00000145362 | ANK2 | ankyrin 2 | -0.45 | 2.778E-03 |
| ENSG00000075826 | SEC31B | SEC31 homolog B, COPII coat complex component | -0.45 | 1.818E-03 |
| ENSG00000122417 | ODF2L | outer dense fiber of sperm tails 2 like | -0.45 | 9.462E-04 |
| ENSG00000119547 | ONECUT2 | one cut homeobox 2 | -0.45 | 6.534E-04 |
| ENSG00000164048 | ZNF589 | zinc finger protein 589 | -0.45 | 3.072E-04 |
| ENSG00000118058 | KMT2A | lysine methyltransferase 2A | -0.45 | 2.522E-04 |
| ENSG00000153914 | SREK1 | splicing regulatory glutamic acid and lysine rich protein 1 | -0.45 | 4.030E-05 |
| ENSG00000167548 | KMT2D | lysine methyltransferase 2D | -0.45 | 1.930E-05 |
| ENSG00000122515 | ZMIZ2 | zinc finger MIZ-type containing 2 | -0.45 | 1.330E-06 |
| ENSG00000100354 | TNRC6B | trinucleotide repeat containing adaptor 6B | -0.44 | 3.376E-02 |
| ENSG00000083097 | DOP1A | DOP1 leucine zipper like protein A | -0.44 | 2.909E-02 |
| ENSG00000266714 | MYO15B | myosin XVB | -0.44 | 2.567E-02 |
| ENSG00000169856 | ONECUT1 | one cut homeobox 1 | -0.44 | 2.282E-02 |
| ENSG00000162836 | ACP6 | acid phosphatase 6, lysophosphatidic | -0.44 | 1.794E-02 |
| ENSG00000163482 | STK36 | serine/threonine kinase 36 | -0.44 | 2.114E-03 |
| ENSG00000198466 | ZNF587 | zinc finger protein 587 | -0.44 | 1.499E-04 |
| ENSG00000066739 | ATG2B | autophagy related 2B | -0.44 | 7.370E-05 |
| ENSG00000134884 | ARGLU1 | arginine and glutamate rich 1 | -0.44 | 2.680E-05 |
| ENSG00000109046 | WSB1 | WD repeat and SOCS box containing 1 | -0.44 | 3.980E-07 |
| ENSG00000179909 | ZNF154 | zinc finger protein 154 | -0.43 | 1.043E-02 |
| ENSG00000052126 | PLEKHA5 | pleckstrin homology domain containing A5 | -0.43 | 8.109E-03 |
| ENSG00000132424 | PNISR | PNN interacting serine and arginine rich protein | -0.43 | 4.097E-03 |
| ENSG00000119707 | RBM25 | RNA binding motif protein 25 | -0.43 | 9.030E-07 |

| | | | | |
|---|---|---|---|---|
| ENSG00000164066 | INTU | inturned planar cell polarity protein | -0.42 | 4.169E-02 |
| ENSG00000185946 | RNPC3 | RNA binding region (RNP1, RRM) containing 3 | -0.42 | 1.896E-02 |
| ENSG00000147454 | SLC25A37 | solute carrier family 25 member 37 | -0.42 | 2.877E-03 |
| ENSG00000120437 | ACAT2 | acetyl-CoA acetyltransferase 2 | -0.42 | 7.390E-05 |
| ENSG00000141449 | GREB1L | GREB1 like retinoic acid receptor coactivator | -0.42 | 8.420E-07 |
| ENSG00000079134 | THOC1 | THO complex 1 | -0.41 | 4.518E-02 |
| ENSG00000143341 | HMCN1 | hemicentin 1 | -0.41 | 2.762E-02 |
| ENSG00000117620 | SLC35A3 | solute carrier family 35 member A3 | -0.41 | 2.664E-02 |
| ENSG00000148690 | FRA10AC1 | FRA10A associated CGG repeat 1 | -0.41 | 8.002E-03 |
| ENSG00000150893 | FREM2 | FRAS1 related extracellular matrix 2 | -0.41 | 7.569E-03 |
| ENSG00000149231 | CCDC82 | coiled-coil domain containing 82 | -0.41 | 1.495E-03 |
| ENSG00000164199 | ADGRV1 | adhesion G protein-coupled receptor V1 | -0.41 | 4.009E-04 |
| ENSG00000184465 | WDR27 | WD repeat domain 27 | -0.41 | 7.730E-05 |
| ENSG00000129003 | VPS13C | vacuolar protein sorting 13 homolog C | -0.41 | 5.630E-06 |
| ENSG00000127481 | UBR4 | ubiquitin protein ligase E3 component n-recognin 4 | -0.41 | 3.780E-10 |
| ENSG00000196705 | ZNF431 | zinc finger protein 431 | -0.4 | 3.286E-02 |
| ENSG00000204104 | TRAF3IP1 | TRAF3 interacting protein 1 | -0.4 | 2.376E-02 |
| ENSG00000131127 | ZNF141 | zinc finger protein 141 | -0.4 | 2.180E-02 |
| ENSG00000198185 | ZNF334 | zinc finger protein 334 | -0.4 | 1.410E-02 |
| ENSG00000138756 | BMP2K | BMP2 inducible kinase | -0.4 | 8.696E-03 |
| ENSG00000169045 | HNRNPH1 | heterogeneous nuclear ribonucleoprotein H1 | -0.4 | 1.598E-03 |
| ENSG00000159086 | PAXBP1 | PAX3 and PAX7 binding protein 1 | -0.4 | 1.232E-03 |
| ENSG00000160145 | KALRN | kalirin RhoGEF kinase | -0.4 | 8.817E-04 |
| ENSG00000112972 | HMGCS1 | 3-hydroxy-3-methylglutaryl-CoA synthase 1 | -0.4 | 6.160E-08 |
| ENSG00000154760 | SLFN13 | schlafen family member 13 | -0.39 | 4.730E-02 |
| ENSG00000131914 | LIN28A | lin-28 homolog A | -0.39 | 3.440E-02 |
| ENSG00000184144 | CNTN2 | contactin 2 | -0.39 | 2.679E-02 |
| ENSG00000205517 | RGL3 | ral guanine nucleotide dissociation stimulator like 3 | -0.39 | 1.975E-02 |
| ENSG00000147421 | HMBOX1 | homeobox containing 1 | -0.39 | 1.741E-02 |
| ENSG00000154358 | OBSCN | obscurin, cytoskeletal calmodulin and titin-interacting RhoGEF | -0.39 | 1.737E-02 |
| ENSG00000135164 | DMTF1 | cyclin D binding myb like transcription factor 1 | -0.39 | 1.680E-03 |
| ENSG00000197969 | VPS13A | vacuolar protein sorting 13 homolog A | -0.39 | 1.659E-03 |
| ENSG00000137504 | CREBZF | CREB/ATF bZIP transcription factor | -0.39 | 6.667E-04 |
| ENSG00000146830 | GIGYF1 | GRB10 interacting GYF protein 1 | -0.39 | 3.014E-04 |
| ENSG00000161996 | WDR90 | WD repeat domain 90 | -0.38 | 3.390E-02 |
| ENSG00000285437 | POLR2J3 | RNA polymerase II subunit J3 | -0.38 | 2.810E-02 |

| | | | | |
|---|---|---|---|---|
| ENSG00000074755 | ZZEF1 | zinc finger ZZ-type and EF-hand domain containing 1 | -0.38 | 1.369E-02 |
| ENSG00000173889 | PHC3 | polyhomeotic homolog 3 | -0.38 | 8.479E-03 |
| ENSG00000007392 | LUC7L | LUC7 like | -0.38 | 3.777E-03 |
| ENSG00000004777 | ARHGAP33 | Rho GTPase activating protein 33 | -0.38 | 3.189E-03 |
| ENSG00000116580 | GON4L | gon-4 like | -0.38 | 2.208E-03 |
| ENSG00000100941 | PNN | pinin, desmosome associated protein | -0.38 | 1.675E-03 |
| ENSG00000105443 | CYTH2 | cytohesin 2 | -0.38 | 2.250E-05 |
| ENSG00000196458 | ZNF605 | zinc finger protein 605 | -0.37 | 3.883E-02 |
| ENSG00000197961 | ZNF121 | zinc finger protein 121 | -0.37 | 2.091E-02 |
| ENSG00000099326 | MZF1 | myeloid zinc finger 1 | -0.37 | 2.060E-02 |
| ENSG00000165699 | TSC1 | TSC complex subunit 1 | -0.37 | 1.967E-02 |
| ENSG00000165115 | KIF27 | kinesin family member 27 | -0.37 | 1.845E-02 |
| ENSG00000258890 | CEP95 | centrosomal protein 95 | -0.37 | 1.655E-02 |
| ENSG00000085224 | ATRX | ATRX chromatin remodeler | -0.37 | 3.984E-03 |
| ENSG00000157796 | WDR19 | WD repeat domain 19 | -0.37 | 8.908E-04 |
| ENSG00000185222 | TCEAL9 | transcription elongation factor A like 9 | -0.37 | 1.070E-05 |
| ENSG00000104722 | NEFM | neurofilament medium | -0.36 | 4.423E-02 |
| ENSG00000126870 | WDR60 | WD repeat domain 60 | -0.36 | 3.972E-02 |
| ENSG00000197323 | TRIM33 | tripartite motif containing 33 | -0.36 | 3.064E-02 |
| ENSG00000188811 | NHLRC3 | NHL repeat containing 3 | -0.36 | 2.902E-02 |
| ENSG00000161010 | MRNIP | MRN complex interacting protein | -0.36 | 1.775E-02 |
| ENSG00000179240 | GVQW3 | GVQW motif containing 3 | -0.36 | 1.717E-02 |
| ENSG00000219481 | NBPF1 | NBPF member 1 | -0.36 | 1.166E-02 |
| ENSG00000005810 | MYCBP2 | MYC binding protein 2 | -0.36 | 1.044E-02 |
| ENSG00000100288 | CHKB | choline kinase beta | -0.36 | 1.043E-02 |
| ENSG00000173166 | RAPH1 | Ras association (RalGDS/AF-6) and pleckstrin homology domains 1 | -0.36 | 8.437E-03 |
| ENSG00000102901 | CENPT | centromere protein T | -0.36 | 8.265E-03 |
| ENSG00000117616 | RSRP1 | arginine and serine rich protein 1 | -0.36 | 4.557E-03 |
| ENSG00000104177 | MYEF2 | myelin expression factor 2 | -0.36 | 2.189E-03 |
| ENSG00000198707 | CEP290 | centrosomal protein 290 | -0.36 | 1.675E-03 |
| ENSG00000138688 | KIAA1109 | KIAA1109 | -0.36 | 1.283E-03 |
| ENSG00000184677 | ZBTB40 | zinc finger and BTB domain containing 40 | -0.35 | 4.395E-02 |
| ENSG00000152104 | PTPN14 | protein tyrosine phosphatase non-receptor type 14 | -0.35 | 3.159E-02 |
| ENSG00000117625 | RCOR3 | REST corepressor 3 | -0.35 | 2.388E-02 |
| ENSG00000197162 | ZNF785 | zinc finger protein 785 | -0.35 | 2.205E-02 |
| ENSG00000183426 | NPIPA1 | nuclear pore complex interacting protein family member A1 | -0.35 | 1.875E-02 |
| ENSG00000110888 | CAPRIN2 | caprin family member 2 | -0.35 | 1.212E-02 |
| ENSG00000047188 | YTHDC2 | YTH domain containing 2 | -0.35 | 4.152E-03 |
| ENSG00000109920 | FNBP4 | formin binding protein 4 | -0.35 | 3.644E-03 |

| | | | | |
|---|---|---|---|---|
| ENSG00000122566 | HNRNPA2B1 | heterogeneous nuclear ribonucleoprotein A2/B1 | -0.34 | 3.796E-02 |
| ENSG00000139718 | SETD1B | SET domain containing 1B, histone lysine methyltransferase | -0.34 | 3.608E-02 |
| ENSG00000185658 | BRWD1 | bromodomain and WD repeat domain containing 1 | -0.34 | 1.362E-02 |
| ENSG00000167978 | SRRM2 | serine/arginine repetitive matrix 2 | -0.34 | 1.266E-02 |
| ENSG00000165434 | PGM2L1 | phosphoglucomutase 2 like 1 | -0.34 | 1.097E-02 |
| ENSG00000139910 | NOVA1 | NOVA alternative splicing regulator 1 | -0.34 | 6.707E-03 |
| ENSG00000080603 | SRCAP | Snf2 related CREBBP activator protein | -0.34 | 9.462E-04 |
| ENSG00000082438 | COBLL1 | cordon-bleu WH2 repeat protein like 1 | -0.33 | 4.960E-02 |
| ENSG00000096093 | EFHC1 | EF-hand domain containing 1 | -0.33 | 4.764E-02 |
| ENSG00000185246 | PRPF39 | pre-mRNA processing factor 39 | -0.33 | 9.782E-03 |
| ENSG00000112159 | MDN1 | midasin AAA ATPase 1 | -0.33 | 9.012E-03 |
| ENSG00000003402 | CFLAR | CASP8 and FADD like apoptosis regulator | -0.33 | 3.645E-03 |
| ENSG00000173230 | GOLGB1 | golgin B1 | -0.33 | 1.675E-03 |
| ENSG00000127603 | MACF1 | microtubule actin crosslinking factor 1 | -0.33 | 1.821E-04 |
| ENSG00000003756 | RBM5 | RNA binding motif protein 5 | -0.33 | 2.250E-05 |
| ENSG00000114127 | XRN1 | 5'-3' exoribonuclease 1 | -0.32 | 4.435E-02 |
| ENSG00000079102 | RUNX1T1 | RUNX1 partner transcriptional co-repressor 1 | -0.32 | 4.011E-02 |
| ENSG00000196440 | ARMCX4 | armadillo repeat containing X-linked 4 | -0.32 | 2.403E-02 |
| ENSG00000166987 | MBD6 | methyl-CpG binding domain protein 6 | -0.32 | 3.582E-03 |
| ENSG00000123636 | BAZ2B | bromodomain adjacent to zinc finger domain 2B | -0.32 | 2.603E-03 |
| ENSG00000151150 | ANK3 | ankyrin 3 | -0.32 | 3.395E-04 |
| ENSG00000011021 | CLCN6 | chloride voltage-gated channel 6 | -0.31 | 3.959E-02 |
| ENSG00000150593 | PDCD4 | programmed cell death 4 | -0.31 | 3.786E-02 |
| ENSG00000115977 | AAK1 | AP2 associated kinase 1 | -0.31 | 2.664E-02 |
| ENSG00000089335 | ZNF302 | zinc finger protein 302 | -0.31 | 1.256E-02 |
| ENSG00000136944 | LMX1B | LIM homeobox transcription factor 1 beta | -0.31 | 1.044E-02 |
| ENSG00000090924 | PLEKHG2 | pleckstrin homology and RhoGEF domain containing G2 | -0.31 | 1.003E-02 |
| ENSG00000067064 | IDI1 | isopentenyl-diphosphate delta isomerase 1 | -0.31 | 8.696E-03 |
| ENSG00000131437 | KIF3A | kinesin family member 3A | -0.31 | 4.149E-03 |
| ENSG00000116754 | SRSF11 | serine and arginine rich splicing factor 11 | -0.31 | 2.726E-03 |
| ENSG00000221978 | CCNL2 | cyclin L2 | -0.31 | 4.040E-05 |

| | | | | |
|---|---|---|---|---|
| ENSG00000177200 | CHD9 | chromodomain helicase DNA binding protein 9 | -0.3 | 4.738E-02 |
| ENSG00000048707 | VPS13D | vacuolar protein sorting 13 homolog D | -0.3 | 4.169E-02 |
| ENSG00000115414 | FN1 | fibronectin 1 | -0.3 | 3.786E-02 |
| ENSG00000078687 | TNRC6C | trinucleotide repeat containing adaptor 6C | -0.3 | 3.128E-02 |
| ENSG00000137343 | ATAT1 | alpha tubulin acetyltransferase 1 | -0.3 | 3.094E-02 |
| ENSG00000157741 | UBN2 | ubinuclein 2 | -0.3 | 3.005E-02 |
| ENSG00000134909 | ARHGAP32 | Rho GTPase activating protein 32 | -0.3 | 1.410E-02 |
| ENSG00000171316 | CHD7 | chromodomain helicase DNA binding protein 7 | -0.3 | 1.266E-02 |
| ENSG00000164050 | PLXNB1 | plexin B1 | -0.3 | 4.498E-03 |
| ENSG00000131051 | RBM39 | RNA binding motif protein 39 | -0.3 | 3.189E-03 |
| ENSG00000148798 | INA | internexin neuronal intermediate filament protein alpha | -0.3 | 2.952E-04 |
| ENSG00000130396 | AFDN | afadin, adherens junction formation factor | -0.3 | 2.301E-04 |
| ENSG00000131013 | PPIL4 | peptidylprolyl isomerase like 4 | -0.29 | 2.746E-02 |
| ENSG00000134899 | ERCC5 | ERCC excision repair 5, endonuclease | -0.29 | 2.403E-02 |
| ENSG00000197603 | CPLANE1 | ciliogenesis and planar polarity effector 1 | -0.29 | 2.210E-02 |
| ENSG00000173575 | CHD2 | chromodomain helicase DNA binding protein 2 | -0.29 | 1.542E-02 |
| ENSG00000160285 | LSS | lanosterol synthase | -0.29 | 1.043E-02 |
| ENSG00000113161 | HMGCR | 3-hydroxy-3-methylglutaryl-CoA reductase | -0.29 | 5.084E-03 |
| ENSG00000103657 | HERC1 | HECT and RLD domain containing E3 ubiquitin protein ligase family member 1 | -0.29 | 2.189E-03 |
| ENSG00000119596 | YLPM1 | YLP motif containing 1 | -0.29 | 1.057E-03 |
| ENSG00000147162 | OGT | O-linked N-acetylglucosamine (GlcNAc) transferase | -0.29 | 2.849E-04 |
| ENSG00000143624 | INTS3 | integrator complex subunit 3 | -0.29 | 2.841E-04 |
| ENSG00000100201 | DDX17 | DEAD-box helicase 17 | -0.29 | 6.230E-05 |
| ENSG00000106344 | RBM28 | RNA binding motif protein 28 | -0.28 | 4.050E-02 |
| ENSG00000099194 | SCD | stearoyl-CoA desaturase | -0.28 | 3.943E-02 |
| ENSG00000109756 | RAPGEF2 | Rap guanine nucleotide exchange factor 2 | -0.28 | 3.440E-02 |
| ENSG00000151320 | AKAP6 | A-kinase anchoring protein 6 | -0.28 | 1.896E-02 |
| ENSG00000189180 | ZNF33A | zinc finger protein 33A | -0.28 | 1.674E-02 |
| ENSG00000125844 | RRBP1 | ribosome binding protein 1 | -0.28 | 1.561E-02 |
| ENSG00000086758 | HUWE1 | HECT, UBA and WWE domain containing E3 ubiquitin protein ligase 1 | -0.28 | 1.185E-02 |
| ENSG00000123384 | LRP1 | LDL receptor related protein 1 | -0.28 | 5.599E-03 |

| | | | | |
|---|---|---|---|---|
| ENSG00000047410 | TPR | translocated promoter region, nuclear basket protein | -0.28 | 4.498E-03 |
| ENSG00000152795 | HNRNPDL | heterogeneous nuclear ribonucleoprotein D like | -0.28 | 3.957E-03 |
| ENSG00000074356 | NCBP3 | nuclear cap binding subunit 3 | -0.28 | 2.726E-03 |
| ENSG00000167522 | ANKRD11 | ankyrin repeat domain 11 | -0.28 | 2.457E-03 |
| ENSG00000158321 | AUTS2 | activator of transcription and developmental regulator AUTS2 | -0.28 | 2.457E-03 |
| ENSG00000107929 | LARP4B | La ribonucleoprotein 4B | -0.28 | 2.217E-03 |
| ENSG00000083857 | FAT1 | FAT atypical cadherin 1 | -0.28 | 2.803E-04 |
| ENSG00000176444 | CLK2 | CDC like kinase 2 | -0.27 | 3.134E-02 |
| ENSG00000013573 | DDX11 | DEAD/H-box helicase 11 | -0.27 | 2.575E-02 |
| ENSG00000134186 | PRPF38B | pre-mRNA processing factor 38B | -0.27 | 2.500E-02 |
| ENSG00000109654 | TRIM2 | tripartite motif containing 2 | -0.27 | 2.372E-02 |
| ENSG00000100393 | EP300 | E1A binding protein p300 | -0.27 | 1.811E-02 |
| ENSG00000137776 | SLTM | SAFB like transcription modulator | -0.27 | 1.655E-02 |
| ENSG00000001630 | CYP51A1 | cytochrome P450 family 51 subfamily A member 1 | -0.27 | 1.003E-02 |
| ENSG00000130164 | LDLR | low density lipoprotein receptor | -0.27 | 8.848E-03 |
| ENSG00000120549 | KIAA1217 | KIAA1217 | -0.27 | 5.500E-03 |
| ENSG00000123200 | ZC3H13 | zinc finger CCCH-type containing 13 | -0.27 | 1.818E-03 |
| ENSG00000173064 | HECTD4 | HECT domain E3 ubiquitin protein ligase 4 | -0.27 | 6.964E-04 |
| ENSG00000176953 | NFATC2IP | nuclear factor of activated T cells 2 interacting protein | -0.26 | 4.549E-02 |
| ENSG00000121390 | PSPC1 | paraspeckle component 1 | -0.26 | 2.800E-02 |
| ENSG00000172915 | NBEA | neurobeachin | -0.26 | 2.741E-02 |
| ENSG00000170004 | CHD3 | chromodomain helicase DNA binding protein 3 | -0.26 | 2.510E-02 |
| ENSG00000119760 | SUPT7L | SPT7 like, STAGA complex subunit gamma | -0.26 | 2.455E-02 |
| ENSG00000118418 | HMGN3 | high mobility group nucleosomal binding domain 3 | -0.26 | 1.691E-02 |
| ENSG00000197535 | MYO5A | myosin VA | -0.26 | 1.627E-02 |
| ENSG00000133858 | ZFC3H1 | zinc finger C3H1-type containing | -0.26 | 1.483E-02 |
| ENSG00000038219 | BOD1L1 | biorientation of chromosomes in cell division 1 like 1 | -0.26 | 1.034E-02 |
| ENSG00000128731 | HERC2 | HECT and RLD domain containing E3 ubiquitin protein ligase 2 | -0.26 | 1.011E-02 |
| ENSG00000117523 | PRRC2C | proline rich coiled-coil 2C | -0.26 | 4.438E-04 |
| ENSG00000124177 | CHD6 | chromodomain helicase DNA binding protein 6 | -0.25 | 4.975E-02 |
| ENSG00000139613 | SMARCC2 | SWI/SNF related, matrix associated, actin dependent regulator of chromatin subfamily c member 2 | -0.25 | 4.169E-02 |
| ENSG00000111011 | RSRC2 | arginine and serine rich coiled-coil 2 | -0.25 | 3.783E-02 |

| | | | | |
|---|---|---|---|---|
| ENSG00000119688 | ABCD4 | ATP binding cassette subfamily D member 4 | -0.25 | 3.322E-02 |
| ENSG00000140836 | ZFHX3 | zinc finger homeobox 3 | -0.25 | 3.128E-02 |
| ENSG00000197102 | DYNC1H1 | dynein cytoplasmic 1 heavy chain 1 | -0.25 | 3.893E-04 |
| ENSG00000196220 | SRGAP3 | SLIT-ROBO Rho GTPase activating protein 3 | -0.24 | 4.599E-02 |
| ENSG00000197119 | SLC25A29 | solute carrier family 25 member 29 | -0.24 | 3.279E-02 |
| ENSG00000107863 | ARHGAP21 | Rho GTPase activating protein 21 | -0.24 | 3.101E-02 |
| ENSG00000104549 | SQLE | squalene epoxidase | -0.24 | 2.927E-02 |
| ENSG00000168916 | ZNF608 | zinc finger protein 608 | -0.24 | 1.549E-02 |
| ENSG00000181222 | POLR2A | RNA polymerase II subunit A | -0.24 | 1.044E-02 |
| ENSG00000075292 | ZNF638 | zinc finger protein 638 | -0.23 | 3.881E-02 |
| ENSG00000005483 | KMT2E | lysine methyltransferase 2E (inactive) | -0.22 | 4.365E-02 |
| ENSG00000160752 | FDPS | farnesyl diphosphate synthase | -0.22 | 4.362E-02 |
| ENSG00000118482 | PHF3 | PHD finger protein 3 | -0.22 | 4.227E-02 |
| ENSG00000155368 | DBI | diazepam binding inhibitor, acyl-CoA binding protein | -0.22 | 3.959E-02 |
| ENSG00000198408 | OGA | O-GlcNAcase | -0.22 | 3.896E-02 |
| ENSG00000064607 | SUGP2 | SURP and G-patch domain containing 2 | -0.22 | 3.591E-02 |
| ENSG00000151835 | SACS | sacsin molecular chaperone | -0.21 | 4.449E-02 |
| ENSG00000117713 | ARID1A | AT-rich interaction domain 1A | -0.21 | 3.769E-02 |
| ENSG00000132780 | NASP | nuclear autoantigenic sperm protein | -0.21 | 3.036E-02 |
| ENSG00000054523 | KIF1B | kinesin family member 1B | -0.2 | 2.111E-02 |
| ENSG00000115306 | SPTBN1 | spectrin beta, non-erythrocytic 1 | -0.2 | 1.699E-02 |
| ENSG00000111605 | CPSF6 | cleavage and polyadenylation specific factor 6 | -0.19 | 3.633E-02 |

List of differently expressed genes comparing mock and SARS-CoV-2 infected iPSC-derived pancreatic cells at Day 3 post infection with adjusted P value < 0.05
